# The natural history of symptomatic COVID-19 in Catalonia, Spain: a multi-state model including 109,367 outpatient diagnoses, 18,019 hospitalisations, and 5,585 COVID-19 deaths among 5,627,520 people

**DOI:** 10.1101/2020.07.13.20152454

**Authors:** Edward Burn, Cristian Tebé, Sergio Fernandez-Bertolin, Maria Aragon, Martina Recalde, Elena Roel, Albert Prats-Uribe, Daniel Prieto-Alhambra, Talita Duarte-Salles

## Abstract

**Background:** The natural history of Coronavirus Disease 2019 (COVID-19) has yet to be fully described, with most previous reports focusing on hospitalised patients. Using linked patient-level data, we set out to describe the associations between age, gender, and comorbidities and the risk of outpatient COVID-19 diagnosis, hospitalisation, and/or related mortality.

**Methods:** A population-based cohort study including all individuals registered in Information System for Research in Primary Care (SIDIAP). SIDIAP includes primary care records covering > 80% of the population of Catalonia, Spain, and was linked to region-wide testing, hospital and mortality records. Outpatient diagnoses of COVID-19, hospitalisations with COVID-19, and deaths with COVID-19 were identified between 1^st^ March and 6^th^ May 2020. A multi-state model was used, with cause-specific Cox survival models estimated for each transition.

**Findings:** A total of 5,627,520 individuals were included. Of these, 109,367 had an outpatient diagnosis of COVID-19, 18,019 were hospitalised with COVID-19, and 5,585 died after either being diagnosed or hospitalised with COVID-19. Half of those who died were not admitted to hospital prior to their death. Risk of a diagnosis with COVID-19 peaked first in middle-age and then again for oldest ages, risk for hospitalisation after diagnosis peaked around 70 years old, with all other risks highest at oldest ages. Male gender was associated with an increased risk for all outcomes other than outpatient diagnosis. The comorbidities studied (autoimmune condition, chronic kidney disease, chronic obstructive pulmonary disease, dementia, heart disease, hyperlipidemia, hypertension, malignant neoplasm, obesity, and type 2 diabetes) were all associated with worse outcomes.

**Interpretation:** There is a continued need to protect those at high risk of poor outcomes, particularly the elderly, from COVID-19 and provide appropriate care for those who develop symptomatic disease. While risks of hospitalisation and death are lower for younger populations, there is a need to limit their role in community transmission. These findings should inform public health strategies, including future vaccination campaigns.

## Introduction

The natural history of Coronavirus disease 2019 (COVID-19) has yet to be well-established. While a wide range of studies have described outcomes among patients hospitalised with COVID-19,^1,2^ these individuals represent only a fraction of those who develop symptomatic disease. Similarly, while outcomes for tested populations have also revealed important insights,^3,4^ COVID-19 testing has often been prioritised on the basis of individuals’ symptoms or perceived risk of outcomes, making inferences about specific risk factors for progression among these populations difficult.^5^

A full description of the natural history of COVID-19 from symptomatic to severe disease is needed. Such a description requires comprehensive patient-level data that captures incident, symptomatic cases from a representative population, with subsequent longitudinal follow-up, and where outcomes such as hospitalisations and mortality, both inside and outside of the hospital setting, can be identified. Moreover, assessing the impact of chronic health conditions on the course of the disease requires comprehensive data on patients’ medical histories. Linked real world data from countries with universal taxpayer-funded primary care-based health systems where general practitioners are the first point of contact for care, and where this role has been maintained in the response to COVID-19, provide a unique opportunity for this purpose.

In Spain, one of the European countries worst hit by the COVID-19 pandemic, primary care has continued to play an important role in the response to the disease. In Catalonia, an autonomous region of Spain with a devolved health system, more than 120,000 outpatient cases of COVID-19 were diagnosed between 15^th^ March and 24^th^ April 2020.^6^ Meanwhile, a nationwide seroprevalence study conducted between 27^th^ April to 11^th^ May, 2020 found the prevalence of IgG antibodies to be around 5% in Spain, and 7% in Catalonia.^7^ Consequently, despite the burden of disease already experienced, there is a need to better understand the features of COVID-19 so as to inform the continued national and global response to it.

Our aim in this study was to describe the associations between age, gender, and comorbidities and risks of COVID-19 diagnosis, hospitalisation with COVID-19, and a COVID-19-related death. The research questions addressed are inferential, assessing the existence of relationships but not the underlying mechanisms or reasons for them.^8^

## Methods

### Study design

This study has been informed by primary care records from Catalonia, Spain, which were linked to COVID-19 test results, hospital records, and mortality data. The resulting database was comprehensive, allowing for the identification of various key events in the progression of COVID-19 over the study period, which began on the 1^st^ March 2020 and ended on the 6^th^ May 2020. A multi-state cohort model provided the framework for analysis, allowing for a systematic consideration of transitions for the general population to diagnosis of COVID-19, hospitalisation with COVID-19, and COVID-19 mortality.

### Study participants, setting and data source

Individual-level routinely-collected primary care data were extracted from the Information System for Research in Primary Care (SIDIAP; www.sidiap.org) database, which captures patient records from approximately 80% of the Catalan population and is representative in geography, age, and gender.^9^ Linkage was made at an individual-level to COVID-19 PT-PCR testing data, hospital data, and regional mortality data.

The entire database has been mapped to the Observational Medical Outcomes Partnership (OMOP) Common Data Model (CDM). This provided a means of structuring the data to a standardised format, and allowed for the application of analytical tools developed by the open-science Observational Health Data Sciences and Informatics (OHDSI) network.^10^

All individuals in SIDIAP as of the 1^st^ March 2020 were identified. Those individuals with at least one year of prior history available, no prior clinical diagnosis or positive test result for COVID-19, and who were not hospitalised on the 1^st^ March 2020 were included in the study. Study follow-up began on the 1^st^ March 2020 and ended on the 6^th^ May 2020 (the last date of available data).

### Variables

Individuals’ age and gender were extracted. Health conditions, using individual’s observed medical history were identified. The Charlson comorbidity index was calculated for individuals using the algorithm previously developed by OHDSI,^11^ and scores were categorised as 0, 1, 2, or 3+. Ten specific health conditions were also extracted, all of interest as potential risk factors for the progression of COVID-19. Autoimmune condition (which included type 1 diabetes, rheumatoid arthritis, psoriasis, psoriatic arthritis, multiple sclerosis, systemic lupus erythematosus, Addison’s disease, Grave’s disease, Sjorgen’s syndrome, Hashimoto thyroiditis, Myasthenia gravis, vasculitis, pernicious anemia, celiac disease, scleroderma, sarcoidosis, ulcerative colitis, and Crohn’s disease), chronic kidney disease, chronic obstructive pulmonary disease (COPD), dementia, heart disease, hyperlipidemia, hypertension, malignant neoplasm excluding non-melanoma skin cancer, type 2 diabetes mellitus were each identified on the basis of diagnosis codes. Obesity was identified either by a diagnosis code, a record of a body mass index measurement between 30 and 60 kg/m2, or a recorded weight between 120 and 200 kg within 5 years of the index date.

Study outcomes included an outpatient clinical diagnosis of COVID-19, a hospitalisation with COVID-19, and death. Outpatient COVID-19 diagnoses were identified on the basis of a compatible clinical code being recorded for COVID-19 disease (ICD-10-CM codes B34.2 “Coronavirus infection, unspecified” and B97.29 “Other coronavirus as the cause of diseases classified elsewhere”). Hospitalisation with COVID-19 was identified as a hospital admission where the individual had a positive PT-PCR test result or a clinical diagnosis of COVID-19 over the 21 days prior to their admission up to the end of their hospital stay. Mortality was identified using region-wide mortality data, and so included both deaths during hospitalisations and in the community.

## Statistical methods

### Multi-state model

A multi-state model provided the framework for the analysis. Multi-state models allow for a consideration of individuals progression to multiple events of interest, extending on competing risk models by also describing transitions to intermediate events.^12^ In the context of COVID-19, clinical diagnoses of the disease and hospitalisations with the disease can be considered as key intermediate events between not being infected (or at least, not having been identified as being so) on one end to death on the other.

The structure of the multi-state model used in this study is shown in Figure 1. There were four states: *general population, outpatient diagnosis with COVID-19, hospitalised with* C*OVID-19*, and *death*. The model is progressive with all individuals beginning in the *general population* state (as described above, individuals included in the study had no prior history of COVID-19 and were not hospitalised on their index date). Six transitions were possible: from *general population* to *outpatient diagnosis with COVID-19*, from *general population* to *hospitalised with COVID-19* (i.e. individuals who did not get a clinical diagnosis prior to hospital admission), from *general population* to *death*, from *outpatient diagnosis with COVID-19* to *hospitalised with COVID-19*, from *outpatient diagnosis with COVID-19* to *death*, and from *hospitalised with COVID-19* to *death*. Given the research objectives, the analyses focused on the five transitions directly related to COVID-19, with the *general population* to *death* used primarily as a means of accounting for the competing risk of death for the study population (although it should be noted that individuals who died with COVID-19, but had not been diagnosed or received a positive test will also be included in this transition).

**Figure 1.**
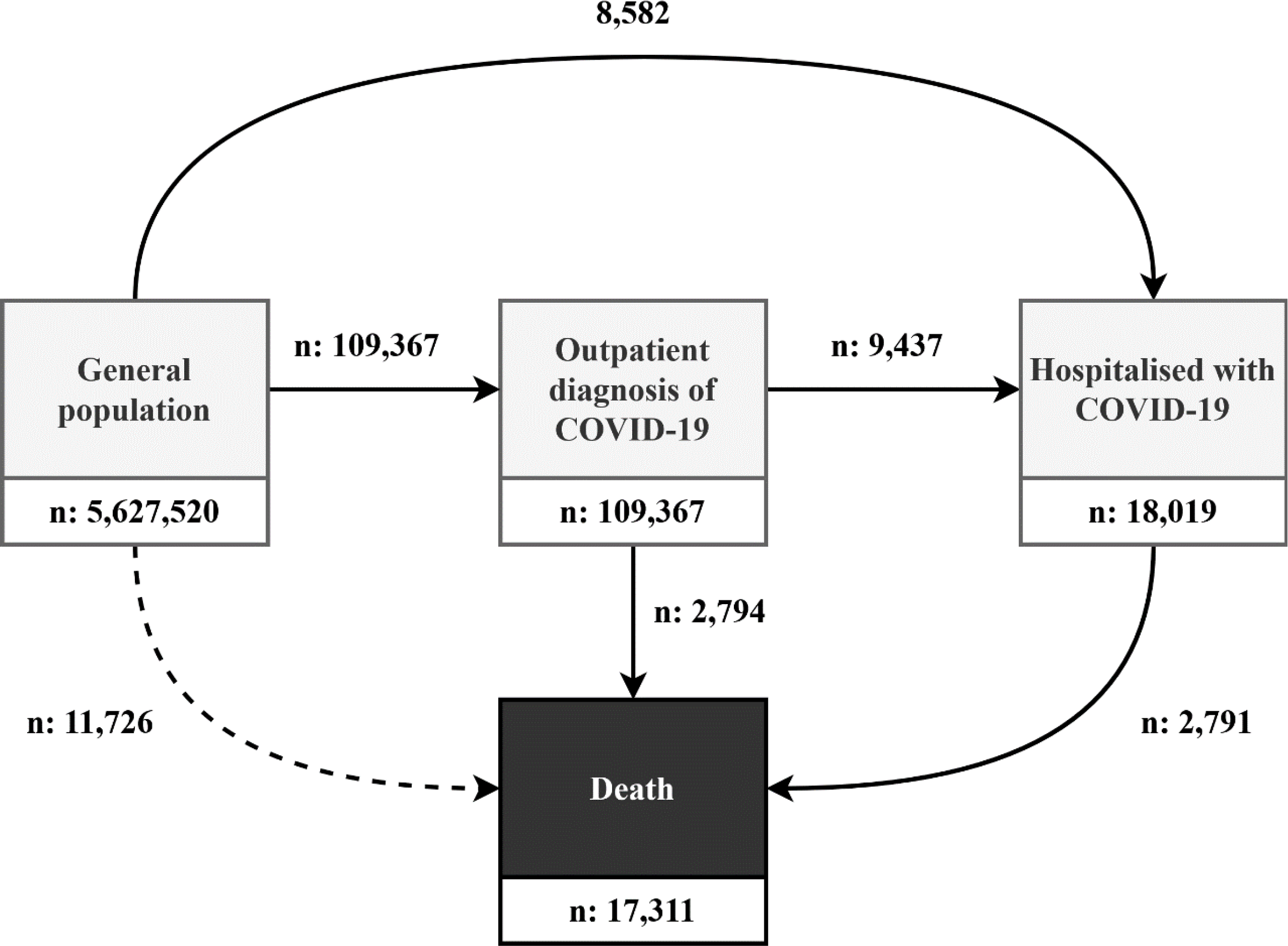
Multi-state model of COVID-19. The entire study population began in the general population state as of the 1^st^ March 2020, with progression through the model possible up to 6^th^ May 2020.

### Describing the association between age and risk of COVID-19 diagnosis, hospitalisation, and death

The relationship between age and the risk of each transition in the multistate model was assessed by estimating cause-specific Cox models, with hazard ratios and 95% confidence intervals (95% CIs) calculated. A non-linear relationship between age and the risk of transitions was considered by fitting age with a polynomial of degree 2 (i.e. quadratic) and with restricted cubic splines (with 3, 4, or 5 knots).^13^ Comparisons between these, and a model where age was assumed to have a linear relationship, were made using Akaike information criterion (AIC) and Bayesian Information Criterion (BIC). Where a more parsimonious model was seen to well-approximate a more complex one, the simpler model was favoured. Models were first estimated with age as the sole explanatory factor, with additional adjustment subsequently made for comorbidities, with the Charlson score and the ten specific conditions of interest described above also included as explanatory factors in the model. To consider whether associations between age and outcomes varied by gender, models were also estimated separately for males and females.

### Describing the association between gender and risk of COVID-19 diagnosis, hospitalisation, and death

The relationship between gender and the risk of each transition in the multistate model was also assessed by estimating cause-specific Cox models. Gender and age were first included as explanatory factors, with non-linearity in age incorporated in the same way as for the previous research objective. Subsequently, as above, adjustment was also made for comorbidities. To consider whether associations between gender and outcomes varied by age, models were also estimated separately for those aged 70 or younger and those aged over 70 (with age assumed to have a linear relationship within each of the strata).

### Describing the association between comorbidities and risk of transitions

The relationship between comorbidities and each transition was assessed by estimating cause-specific Cox models. Models were estimated for each comorbidity of interest separately, controlling for age and gender and with non-linearity in age incorporated in the same way as described above. To consider whether associations between comorbidities and outcomes varied by age and gender, models were subsequently estimated stratifying by both age (70 or younger or above 70) and gender, with age also included as an explanatory factor within each of the strata as a linear term.

### Software used and code availability

The mapping of source SIDIAP data to the OMOP CDM was facilitated by various open-source OHDSI software, which included usagi,^14^ to help define mappings from source codes to the standard concepts used in the CDM, and Achilles,^15^ to help assess data quality after mapping. Data analysis was performed in R version 4.0.0. The R packages used in the analysis included OHDSI CohortDiagnostics,^16^ numerous tidyverse packages,^17^ mstate,^18^ and rms.^19^ The analytic code used in this study is freely available at https://github.com/SIDIAP/MultiStateCovid-19. Cohort definitions were adapted from those used in the OHDSI Seek COVER study and the OHDSI CHARYBDIS project.^20,21^ A web application which summarises the cohort definitions used is available at https://livedataoxford.shinyapps.io/MultiStateCovidCohorts/.

## Results

### Study participants and observed outcomes

A total of 5,627,520 participants were included in the study. Based on the study eligibility criteria described above, 169,577 individuals were excluded due to a lack of a year of prior history, fewer than 5 people for a previous positive COVID-19 test result, 312 for a prior clinical diagnosis of COVID-19, 19 for a prior hospitalisation with COVID-19, and 1,306 who were hospitalised on the 1^st^ March 2020). The characteristics of the included study population are summarised in Table 1, a histogram of the age distribution can be seen in Figure A1, and a summary of comorbidities are shown in Figure 2.

**Table 1.** Patient characteristics of the study population and by multi-state transition

|  | General population | From general population |  |  | From diagnosed with COVID-19 | From hospitalised with COVID-19 |  |
| --- | --- | --- | --- | --- | --- | --- | --- |
|  |  | To diagnosed with COVID-19 | To hospitalised with COVID-19 | To death | To hospitalised with COVID-19 | To death | To death |
| n | 5,627,520 | 109,367 | 8,582 | 11,726 | 9,437 | 2,794 | 2,791 |
| Age (median [IQR]) | 44 [25.0, 60.0] | 47 [36.0, 61.0] | 72 [58.2, 81.0] | 85 [76.0, 90.0] | 61 [50.0, 75.0] | 87 [81.0, 91.0] | 81 [73.0, 86.5] |
| Age group (%) |  |  |  |  |  |  |  |
| Under 18 | 967,227 (17.2) | 4,547 (4.2) | 34 (0.4) | 9 (0.1) | 40 (0.4) | <5 | <5 |
| 18 to 39 | 1,438,732 (25.6) | 30,640 (28.0) | 432 (5.0) | 80 (0.7) | 857 (9.1) | 5 (0.2) | 11 (0.4) |
| 40 to 59 | 1,788,832 (31.8) | 44,803 (41.0) | 1,815 (21.1) | 658 (5.6) | 3,477 (36.8) | 63 (2.3) | 143 (5.1) |
| 60 to 69 | 617,929 (11.0) | 10,661 (9.7) | 1,590 (18.5) | 1,034 (8.8) | 1,708 (18.1) | 136 (4.9) | 296 (10.6) |
| 70 to 79 | 474,369 (8.4) | 7,163 (6.5) | 2,252 (26.2) | 1,999 (17.0) | 1,702 (18.0) | 421 (15.1) | 840 (30.1) |
| 80 or older | 340,431 (6.0) | 11,553 (10.6) | 2,459 (28.7) | 7,946 (67.8) | 1,653 (17.5) | 2,169 (77.6) | 1,500 (53.7) |
| Gender: female (%) | 2,859,274 (50.8) | 64,473 (59.0) | 3,770 (43.9) | 5,932 (50.6) | 4,357 (46.2) | 1,625 (58.2) | 1,122 (40.2) |
| Charlson comorbidity index (%) |  |  |  |  |  |  |  |
| 0 | 4,572,265<br>(81.2) | 81,833 (74.8) | 3,750 (43.7) | 1,672<br>(14.3) | 5,513 (58.4) | 391<br>(14.0) | 604 (21.6) |
| 1 | 410,497<br>(7.3) | 10,708 (9.8) | 1,156 (13.5) | 1,724<br>(14.7) | 1,157 (12.3) | 566<br>(20.3) | 447 (16.0) |
| 2 | 363,057<br>(6.5) | 8,211 (7.5) | 1,518 (17.7) | 2,398<br>(20.5) | 1,266 (13.4) | 507<br>(18.1) | 572 (20.5) |
| 3+ | 281,701<br>(5.0) | 8,615 (7.9) | 2,158 (25.1) | 5,932<br>(50.6) | 1,501 (15.9) | 1,330<br>(47.6) | 1,168 (41.8) |
| Autoimmune<br>condition (%) | 277,450<br>(4.9) | 6,936 (6.3) | 871 (10.1) | 1,320<br>(11.3) | 786 (8.3) | 291<br>(10.4) | 340 (12.2) |
| Chronic kidney<br>disease (%) | 211,842<br>(3.8) | 5,843 (5.3) | 1,548 (18.0) | 3,844<br>(32.8) | 1,058 (11.2) | 955<br>(34.2) | 858 (30.7) |
| COPD (%) | 122,745<br>(2.2) | 2,889 (2.6) | 801 (9.3) | 1,488<br>(12.7) | 515 (5.5) | 280<br>(10.0) | 379 (13.6) |
| Dementia (%) | 57,310 (1.0) | 4,795 (4.4) | 598 (7.0) | 2,894<br>(24.7) | 486 (5.1) | 1,108<br>(39.7) | 416 (14.9) |
| Heart disease (%) | 547,795<br>(9.7) | 13,540 (12.4) | 2,846 (33.2) | 5,865<br>(50.0) | 2,132 (22.6) | 1,264<br>(45.2) | 1,371 (49.1) |
| Hyperlipidemia<br>(%) | 520,837<br>(9.3) | 11,888 (10.9) | 1,590 (18.5) | 1,630<br>(13.9) | 1,599 (16.9) | 415<br>(14.9) | 523 (18.7) |
| Hypertension (%) | 703,167<br>(12.5) | 16,400 (15.0) | 2,609 (30.4) | 3,971<br>(33.9) | 2,412 (25.6) | 970<br>(34.7) | 969 (34.7) |
| Malignant<br>neoplasm (%) | 297,409<br>(5.3) | 6,656 (6.1) | 1,405 (16.4) | 3,818<br>(32.6) | 1,075 (11.4) | 626<br>(22.4) | 691 (24.8) |
| Obesity (%) | 937,593<br>(16.7) | 22,493 (20.6) | 3,384 (39.4) | 3,251<br>(27.7) | 3,286 (34.8) | 733<br>(26.2) | 1,118 (40.1) |
| Type 2 Diabetes<br>Mellitus (%) | 324,944<br>(5.8) | 7,427 (6.8) | 1,773 (20.7) | 2,684<br>(22.9) | 1,422 (15.1) | 637<br>(22.8) | 734 (26.3) |

COPD: chronic obstructive pulmonary disease; IQR: interquartile range.

**Figure 2.**
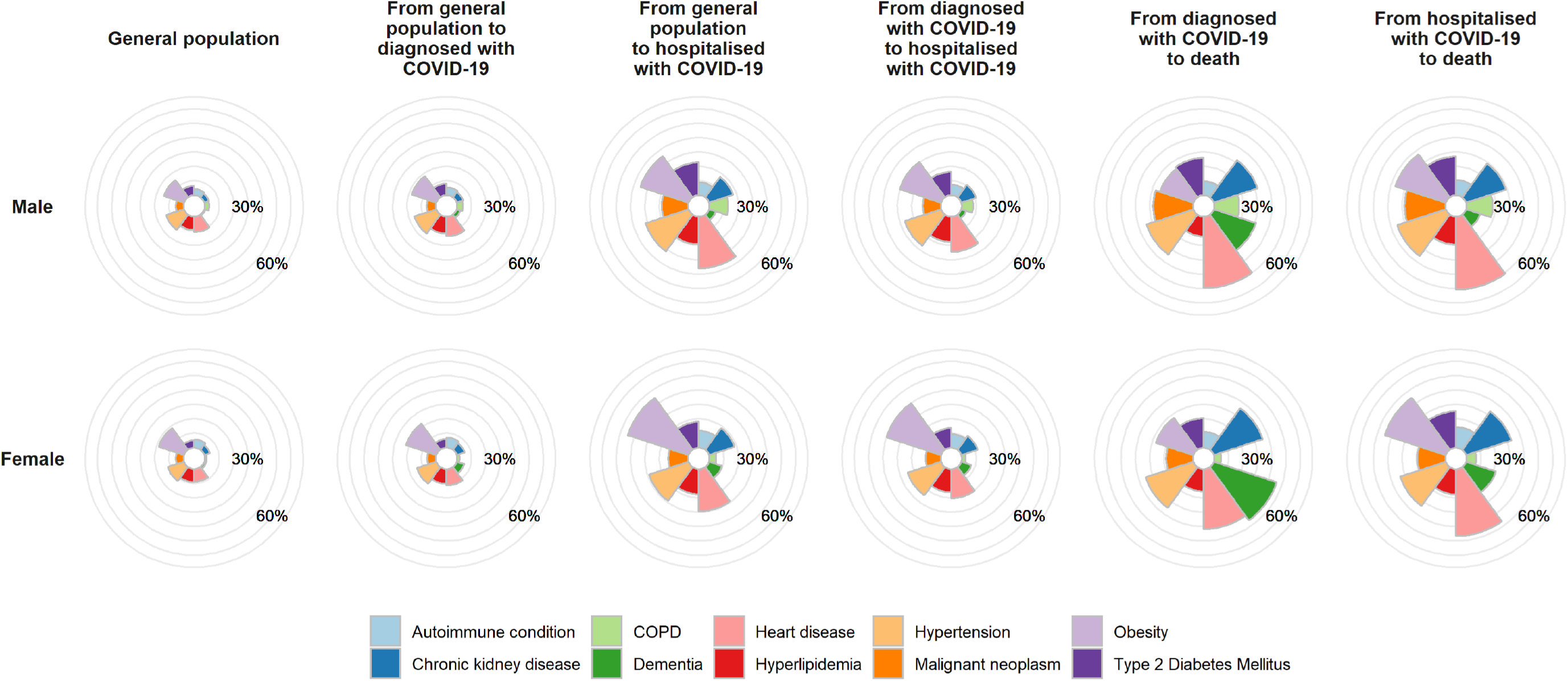
Prevalence of comorbidities in COVID-19 affected and general population. The prevalence of comorbidities of interest are shown here for the study population as a whole, and for the individuals who made each of the transitions in the multi-state model

Out of the included study population, 109,367 went on to have an outpatient diagnosis of COVID-19 (67-day cumulative incidence: 1.94%). Of those diagnosed in outpatient settings, 2,794 died with COVID-19 without being hospitalised (45-day cumulative incidence: 3.11%). In total, 18,019 individuals had a hospitalisation with COVID-19, with 9,437 of them having previously had an outpatient diagnosis (45-day cumulative incidence: 9.03%). Of those hospitalised, 2,791 had died with COVID-19 by the end of follow-up (45-day cumulative incidence: 19.03%), see Figure 1 and Table 2. An extensive summary of the observed data can be seen alongside the study code at https://github.com/SIDIAP/MultiStateCovid-19.

**Table 2.**
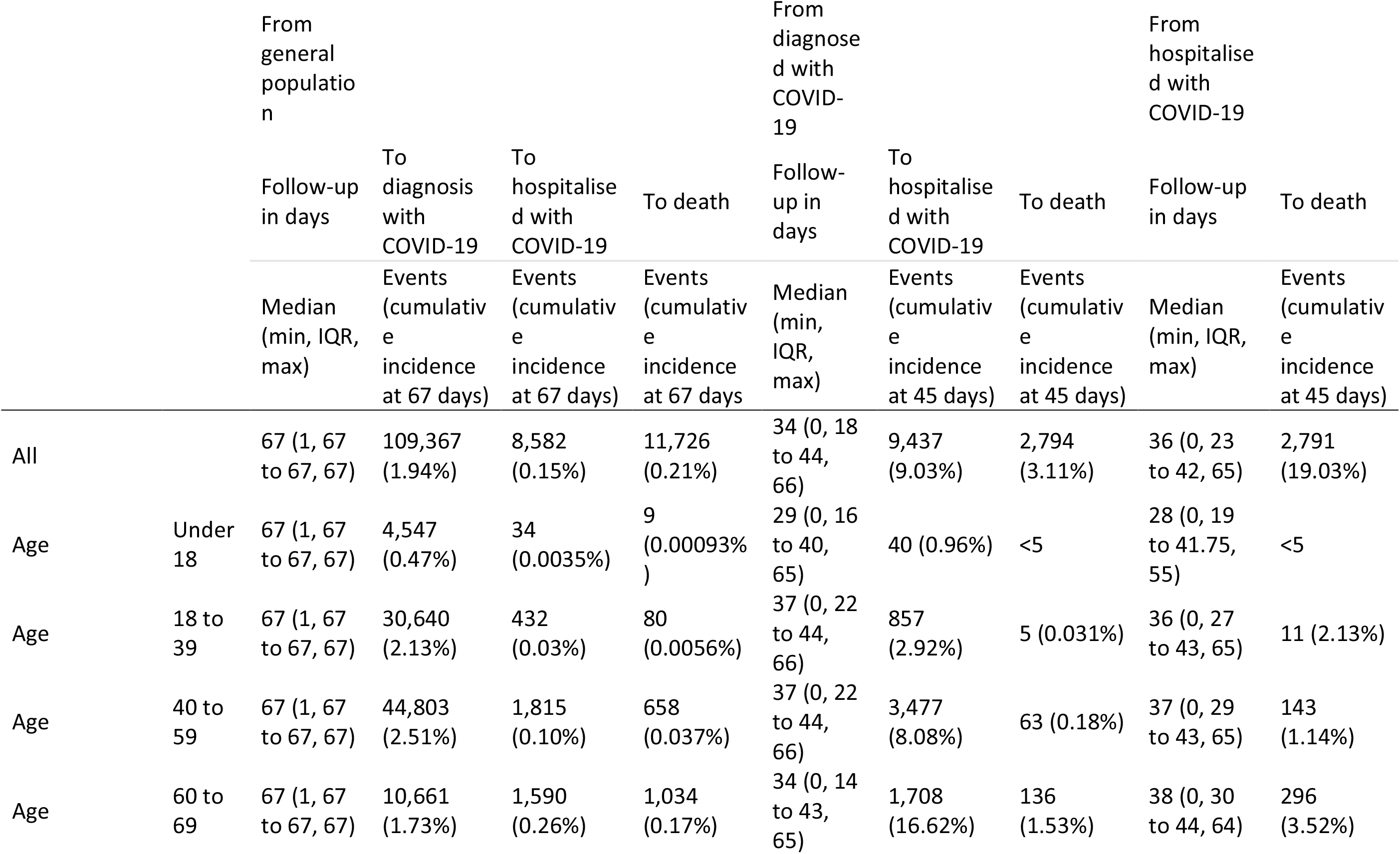
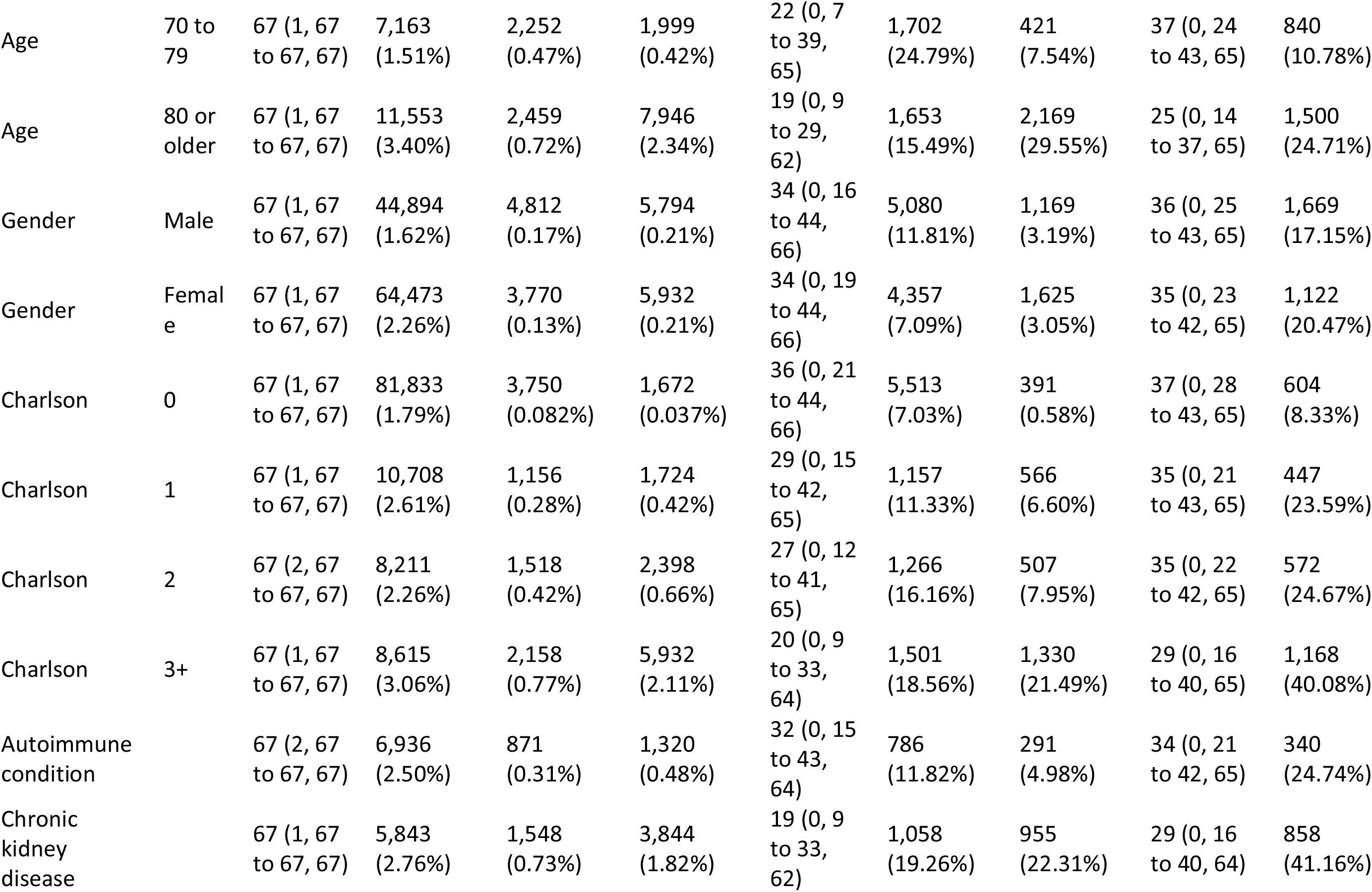
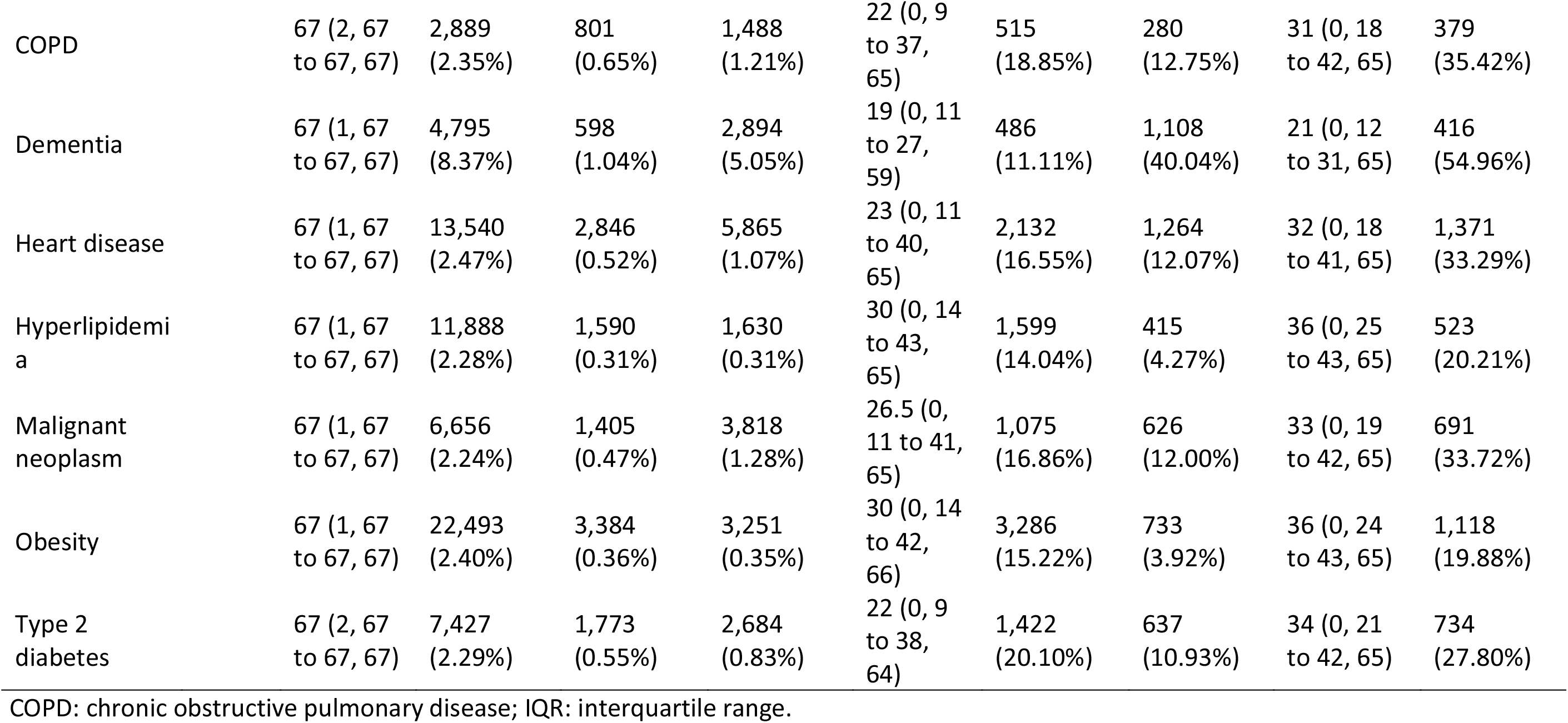
Time at risk and observed outcomes by state populations

### The association between age and risk of COVID-19 diagnosis, hospitalisation, and death

Age profiles varied by transition, see Table 1. While the median age of the study population as a whole was 44 (interquartile range: 25 to 60), those diagnosed with COVID-19 were 47 (IQR: 36 to 61), and those hospitalised without an outpatient diagnosis were 72 (58 to 81). Individuals hospitalised after a diagnosis of COVID-19 were on average 61 (50 to 75) years old, and those who died after being diagnosed with COVID-19 (but who were not admitted to hospital beforehand) had an average age of 87 (81 to 91). Individuals who died after being hospitalised had an average age of 81 (73 to 87).

Estimated hazard ratios for age are shown in Figure 3 and summarised in full detail in Appendix Table A1 to A5. A non-linear relationship can be seen for outpatient diagnosis with COVID-19, with risks peaking first among those aged around 45 and then again at oldest ages. Relative to a reference age of 65, estimated hazard ratios were 0.69 (95% confidence interval: 0.68 to 0.70) for a 20-year-old, 1.62 (1.59 to 1.65) for a 45-year-old, and 2.13 (2.07 to 2.18) for a 90-year-old. Risk of hospitalisation with COVID-19 after an outpatient diagnosis of COVID-19 peaked at around age 70, with estimated hazard ratios of 0.24 (0.22 to 0.25) for a 45-year-old, 1.10 (1.09 to 1.11) for a 70-year-old, and 0.70 (0.65 to 0.74) for a 90-year-old, all relative to a reference age of 65 years old.

**Figure 3.**
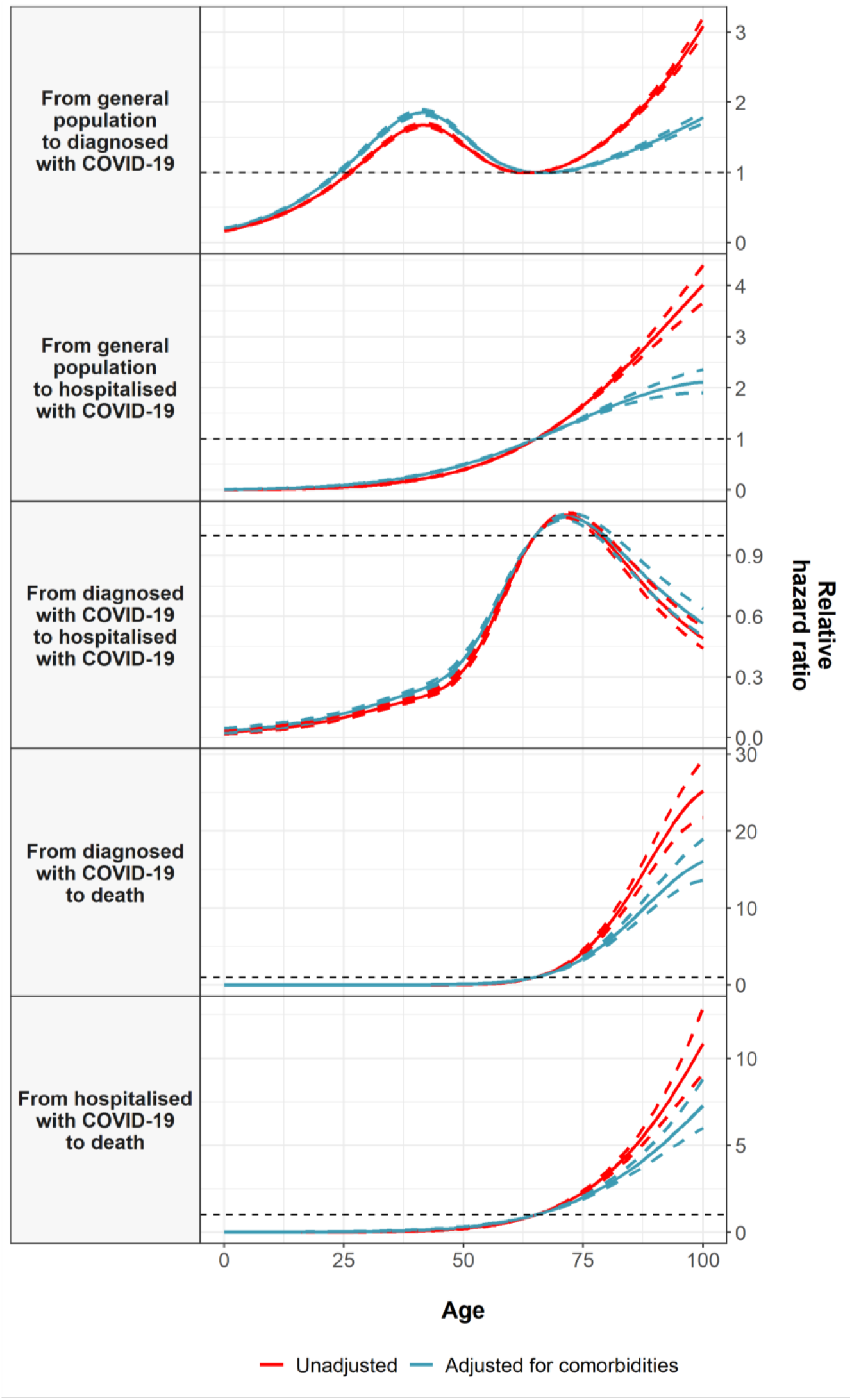
Age and COVID-19 transitions. Estimated hazard ratios for age (relative to a reference age of 65) from cause-specific Cox models for each transition. Dotted lines represent 95% confidence intervals.

Older age was associated with an increased risk of hospitalisation with COVID-19 without a prior diagnosis, death after being hospitalised with COVID-19, and, in particular, death after being diagnosed but without being hospitalised. An age of 90 years old was associated with hazard ratios, relative to 65 years old, of 3.00 (2.84 to 3.16), 6.41 (5.84 to 7.04), and 17.17 (15.55 to 18.97) for each of these, respectively. For an age of 20, these relative hazard ratios were all estimated to be less than 0.01. Adjustment for comorbidities attenuated the associations for older ages to some degree, but not in their entirety. Models estimated separately by gender gave broadly comparable results, see Appendix Tables A1 to A5 and Appendix Figure A2.

### The association between gender and risk of COVID-19 diagnosis, hospitalisation, and death

Compared to the study population as a whole, which was 51% female, a greater proportion of females were diagnosed with COVID-19 (59%), but more males (56%) transitioned directly to being hospitalised with COVID-19 without a previous outpatient diagnosis. Of those diagnosed, more males (54%) were subsequently hospitalised, but more females (58%) died without having been hospitalised. Of those hospitalised, 60% of those who died were male, see Table 1.

After adjustment for age, male gender was associated with a reduced risk of outpatient COVID-19 diagnosis (hazard ratio: 0.74 [95% CI: 0.73 to 0.74]). Conversely, male gender was associated with an increased risk of all other transitions and hence with more severe forms of COVID-19 disease (Figure 4). Hazard ratios for males were 1.61 (1.55 to 1.69) for hospitalisation with COVID-19 without outpatient diagnosis, 1.82 (1.75 to 1.90) for hospitalisation after outpatient diagnosis of COVID-19, 1.75 (1.62 to 1.89) for death after outpatient diagnosis of COVID-19, and 1.41 (1.31 to 1.52) for death after hospitalisation with COVID-19. Further adjustment for comorbidities made little difference to these estimates, see Figure 4. Estimates were also broadly consistent across age-stratified models, although female gender was associated with an even greater increase in risk for the transition from diagnosed with COVID-19 to death for those aged 70 years old or younger, see Appendix Table A5 and Appendix Figure A3.

**Figure 4.**
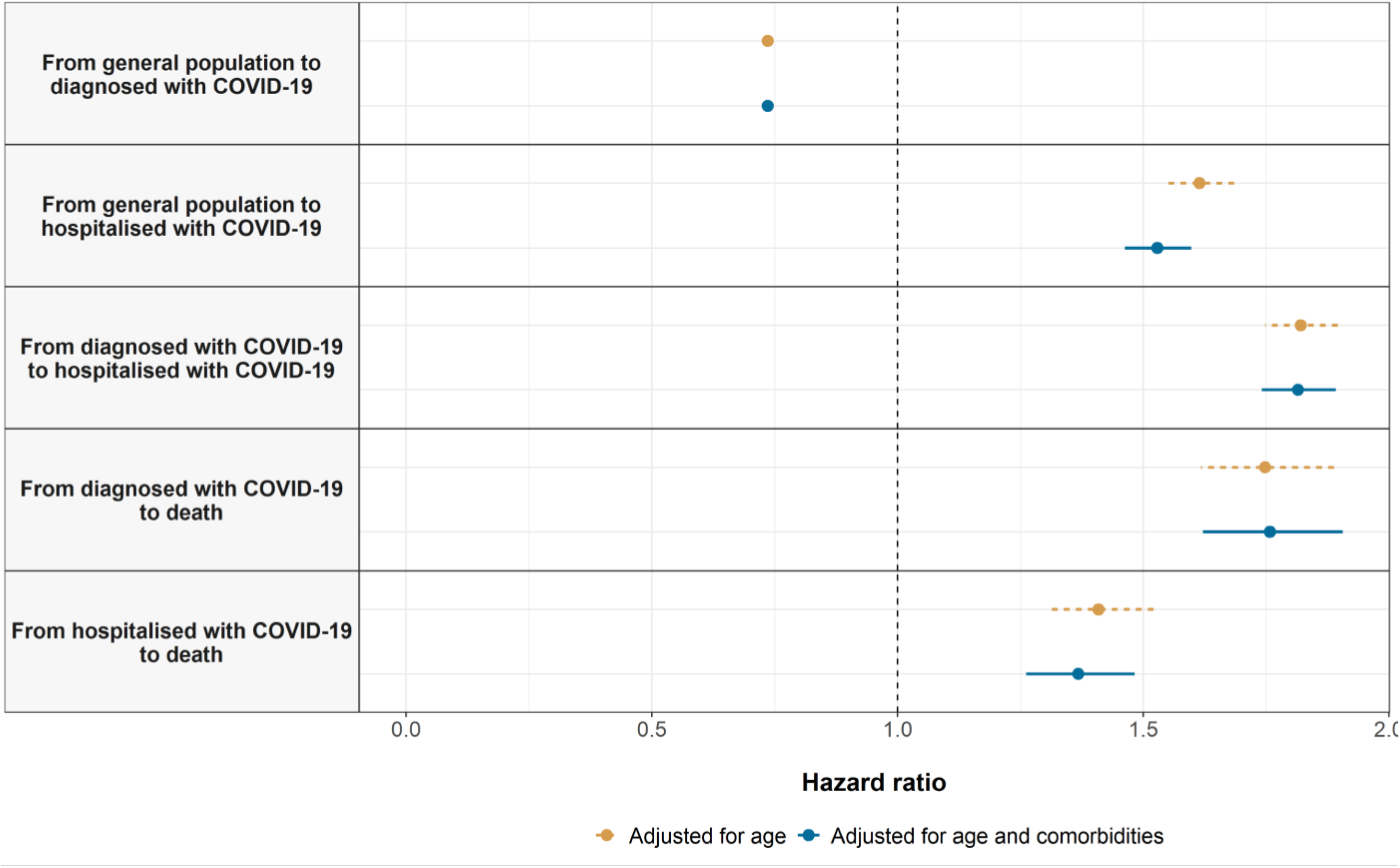
Gender and COVID-19 transitions. Estimated hazard ratios for male gender (relative to female) from cause-specific Cox models for each transition. Points give estimated hazard ratios, with lines representing 95% confidence intervals.

**Figure 5.**
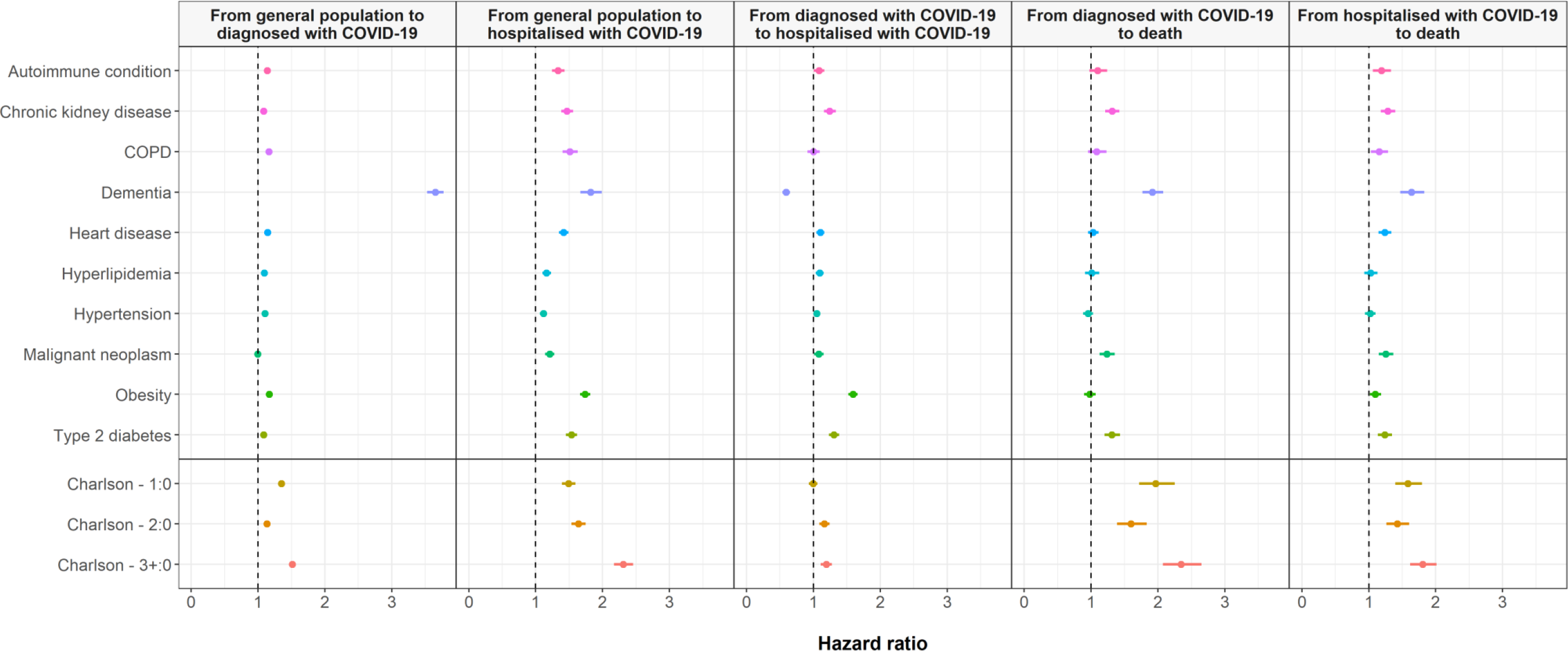
Comorbidities and COVID-19 transitions. Estimated hazard ratios for comorbidities of interest from cause-specific Cox models for each transition. Points give estimated hazard ratios, with lines representing 95% confidence intervals. Models were estimated separately for each comorbidity of interest, with adjustment for age and gender.

### The association between comorbidities and risk of outpatient COVID-19 diagnosis, hospitalisation, and death

Comorbidity profiles varied across different stages of the natural history of COVID-19, see Table 1 and Figure 2. The highest proportion of otherwise healthy participants (Charlson index score of 0) was seen among those diagnosed with COVID-19 in outpatient settings (75%) and lowest amongst those who died after an outpatient diagnosis without hospital admission (14%). While, for example, the prevalence of chronic kidney disease, COPD, obesity, and type 2 diabetes were 4%, 2%, 17%, and 6%, respectively, among the study population as a whole, the prevalence of these conditions was 31%, 14%, 40%, and 26% among those who died after being hospitalised with COVID-19. Of those individuals that died after diagnosis of COVID-19 without have being hospitalised, 40% had dementia (see Table 2). Indeed, among women who died after diagnosis of COVID-19 without have being hospitalised, close to 60% had dementia (see Figure 2).

After accounting for age and gender, dementia had the strongest association with risk of outpatient diagnosis with COVID-19 (hazard ratio: 3.65 [95%CI: 3.53 to 3.77]), see Figure 4. Aside from malignant neoplasm, all other comorbidities were associated with increased risks of outpatient diagnosis, with estimated hazard ratios between 1.08 (1.05 to 1.12) for chronic kidney disease to 1.16 (1.12 to 1.21) for COPD. Similarly, all conditions were associated with an increased risk of hospital admission for COVID-19 without a previous outpatient diagnosis, with obesity (hazard ratio: 1.74 [1.66 to 1.82]) and dementia (1.82 [1.67 to 1.99]) associated with the greatest excess risks.

Dementia was associated with a reduced risk of hospitalisation after diagnosis, but an increased risk of death without hospitalisation, with age and gender adjusted hazard ratios of 0.59 (0.54 to 0.65) and 1.92 (1.77 to 2.08) respectively. Aside from COPD where no association was seen, all other conditions were associated with an increased risk of hospitalisation after outpatient diagnosis, ranging from a hazard ratio of 1.05 (1.00 to 1.10) for hypertension to 1.59 (1.52 to 1.66) for obesity.

While fewer conditions appeared associated with an increased risk of death following outpatient diagnosis, cancer had a hazard ratio of 1.24 (1.13 to 1.35), chronic kidney disease had one of 1.31 (1.21 to 1.42), and type 2 diabetes one of 1.31 (1.20 to 1.43). Among those hospitalised, while little difference in outcomes was seen for those with hyperlipidaemia and hypertension after adjusting for age and gender, other conditions were associated with increased risks with hazard ratios ranging from 1.10 (1.02 to 1.18) for obesity to 1.64 (1.47 to 1.83) for dementia.

The association between the Charlson comorbidity index and transitions was most clearly seen for the risks of hospitalisation without a previous diagnosis and of death following an outpatient diagnosis with COVID-19 without hospitalisation. A score of three or more was, relative to a score of zero, associated with age and gender adjusted hazard ratios of 2.31 (2.17 to 2.46) and 2.34 (2.07 to 2.65) respectively.

While stratified analyses by age and gender generally revealed broadly consistent estimates, various comorbidities were associated with an even greater increase in risk for younger, and particularly female, study participants for a number of transitions, see Appendix Table A6 and Appendix Figure A4 and A5. Obesity, for example, was associated with hazard ratios of 2.67 (2.42 to 2.94) and 2.10 (1.95 to 2.27) for hospitalisation from general population and hospitalisation after a diagnosis with COVID-19.

## Discussion

### Summary of key results

In this large cohort study, primary care data from 5,627,520 individuals with linked COVID-19 testing, hospitalisation, and mortality data has allowed for a detailed characterisation of the natural history of symptomatic COVID-19 as seen during the peak of the epidemic in Spain. Over 100,000 outpatient COVID-19 diagnoses, more than 18,000 hospitalisations with COVID-19, and 5,500 related deaths were observed between 1^st^ March and 6^th^ May. Of these deaths, half were among individuals diagnosed with COVID-19 in outpatient settings but not admitted to hospital.

Older age was consistently associated with increased risks of hospitalisation and mortality, most dramatically for risk of death following an outpatient diagnosis of COVID-19 without a subsequent hospital admission. There was though a notable peak in risk of outpatient diagnosis for people in middle-age. A differential association was seen with gender; while women were at increased risk of outpatient diagnosis, men were consistently at higher risk of hospitalisation and death. Finally, the specific comorbidities studies (autoimmune condition, chronic kidney disease, chronic obstructive pulmonary disease, dementia, heart disease, hyperlipidemia, hypertension, malignant neoplasm, obesity, and type 2 diabetes), and a composite comorbidity index (Charlson Comorbidity Index) were associated with worse outcomes. In a number of cases the increase in risk associated with comorbidities appeared to be particularly pronounced for women aged under 70.

### Findings in context

Older age has consistently been found to be a risk factor for worse outcomes in COVID-19. It has been associated with an increased risk of hospitalisation after testing positive for COVID-19,^3,4^ worse outcomes among those hospitalised,^1,2^ and COVID-19-related mortality among the general population.^22^ Similarly, in our study we find age to be associated with poorer outcomes. Our findings suggest that not only is age associated with increased disease severity, but that there was also likely inequitable access to care during the height of the pandemic in Spain. The increase in risk for age was most strongly seen for the transition from diagnosis to death (with no admission to hospital seen between the two), while the risk of hospitalisation itself was also seen to fall at oldest ages. This may reflect rationing of health care resources, with younger patients likely prioritised for the receipt of hospital care, and intensive services if admitted. With half of the deaths in this study observed among those not hospitalised after diagnosis, who had an average age of 87, it is of uptmost importance that similar individuals should be given appropriate access to care in the future. Our results also provide further insights on community transmission of COVID-19. A peak in diagnosis of COVID-19 was seen among individuals around the age of 45 years old. While this may to some degree reflect differences in health seeking behaviour across age groups, it likely also reflects differences in infection rates across age groups. A large and well-conducted study in Spain found that seroprevalence rose until plateauing at around age of 45 based on point-care-tests, but that seroprevalence was lower for those older than 85 compared to younger adults given immunoassay results.^7^ Meanwhile, a seroprevalence study in Geneva found that those aged between 20 and 49 had the highest likelihood of being seropositive.^23^ Consequently, while seemingly facing lower risks of poor outcomes if infected, younger age groups are being infected and appear to contribute to the spread of the virus in the community.

Male gender has also been associated with worse prognosis in COVID-19, with increased risks for both hospitalisation among those tested,^3^ and worse outcomes among those hospitalised.^2,3^ Our data are compatible with previous literature, with risks of admission and mortality all increased for males. However, we found the opposite effect for outpatient diagnosis, with women being at increased risk of diagnosis with COVID-19 in the community. While this finding contrasts with two UK studies, which reported a higher risk of testing positive for SARS-CoV-2 among men,^24,25^ it is congruent with the findings from a study from China that reported a higher attack rate among women.^26^ In Spain, seroprevalence was seen to be similar for males and females.^27^ Further research is therefore needed to understand this finding, which could be explained by differences in exposure and/or vulnerability to severe acute respiratory syndrome coronavirus 2 (SARS-CoV-2), or differences in health seeking behaviour between men and women.

Comorbidities have been associated with worse outcomes in COVID-19 in this study. In line with our findings, those with a higher Charlson score testing positive for COVID-19 have been previously seen to have an increased risk of hospitalisation and death.^4^

Dementia was seen in our study to be associated with a much-increased risk of diagnosis with COVID-19. This pattern likely reflects both outbreaks of COVID-19 in nursing homes, but also increased surveillance with nursing homes systematically screened for COVID-19 in Barcelona.^28^ For those diagnosed, dementia was associated with a reduced risk of hospitalisation, but an increased risk of death without hospitalisation. This may reflect the failure to provide and seek prompt care in these settings, but also difficulties in accessing appropriate hospital care for residents.

Other comorbidities were seen to be associated with worse outcomes, which is in accordance with previous findings on outcomes for individuals with chronic kidney disease, COPD, heart disease, hyperlipidemia, hypertension, cancer, obesity and type 2 diabetes mellitus.^2–4,24,25^ Our findings have also generally shown the effect of type 2 diabetes, obesity, and chronic kidney disease to be most pronounced among women under 70. Fewer studies have assessed the relationship between autoimmune conditions and outcomes. As with other comorbidities, we found autoimmune conditions to be associated with an increased risk of poorer outcomes in COVID-19. Further research is though certainly merited to consider each of these conditions in turn and consider the impact of other factors such as these individuals’ medication use.

### Study limitations

Our study is informed by routinely-collected health care data with various interactions between individuals and the health system identified, covering outpatient diagnoses, COVID-19 testing, and hospitalisations, and with linked mortality data. This though unavoidably misses health outcomes experienced by individuals that do not lead to any interaction with the health care system, with both asymptomatic individuals and a sizeable proportion of mild symptomatic cases unlikely to be seen. Some of the clinical diagnoses observed in the study may also represent false positives, given the uncertainty surrounding the presentation of the disease during the study period. We did not require clinical diagnoses to be confirmed by the presence of a positive PT-PCR test given the limited amount of testing being performed during the study period. Deaths from COVID-19 where individuals were not tested or diagnosed beforehand will also not have been identified.

While providing a broad picture of clinical trajectories, each of the studied states and transitions can, and should, be considered in further detail. Analyses of the provision of intensive care admissions during hospitalisation is one such example, but would require further data with sufficient granularity on inpatient treatment which was beyond the scope of this study. Similarly, the set of comorbidities considered here are only a subset of the myriad set of conditions that are of interest when considering potential risk factors for COVID-19. Our classifications are broad, and we have not attempted to split conditions by severity or concurrent medication use.

The purpose of this study was descriptive in nature. The aim was not prediction, nor was it causal inference. In particular, it should be noted that associations between specific comorbidities and outcomes do not necessarily reflect a causal relationship. Assessing whether a particular chronic condition is the cause worse outcomes in COVID-19 will require further consideration of, and accounting for, relevant confounding factors.

## Conclusion

In this descriptive study we provide a comprehensive overview of the natural history of symptomatic COVID-19 in Catalonia, Spain. The findings from this study can help inform the continued response to COVID-19 both in Spain and elsewhere. Our research has helped to reveal a clear need to protect at risk populations, particularly the elderly, whilst also considering middle-age populations at a particularly high risk of milder infections and therefore likely key in the community transmission of the disease. Future public health strategies, such as potential lockdowns, shielding, and vaccination campaigns, should take these findings into consideration.

## Data Availability

In accordance with current European and national law, the data used in this study is only available for the researchers participating in this study. Thus, we are not allowed to distribute or make publicly available the data to other parties. However, researchers from public institutions can request data from SIDIAP if they comply with certain requirements. Further information is available online (https://www.sidiap.org/index.php/menu-solicitudes-en/application-proccedure) or by contacting Anna Moleras.

## Acknowledgements

We would like to acknowledge the patients who suffered from or died of this devastating disease, and their families and carers. We would also like to thank the healthcare professionals involved in the management of COVID-19 during these challenging times, from primary care to intensive care units in the Catalan healthcare system.

Voldríem reconéixer i tindre un record per tots els pacients que han patit i els què han mort per la COVID-19. Volem també agrair tots els professionals sanitaris que han diagnosticat i tractat aquesta malaltia al sistema català de salut, des dels centres d’atenció primària fins les unitats de cures intensives.

Queremos reconocer y recordar a todos los pacientes que han sufrido y muerto por la COVID-19. Queremos también agradecer a todos los profesionales sanitarios que han diagnosticado y tratado esta enfermedad en el sistema catalán de salud y en España, desde los centros de salud hasta las unidades de cuidados intensivos.

The analysis has made use of a range of open-source, free available tools provided by the OHDSI community. The phenotypes used in this study were developed, or adapted from, work performed during the OHDSI COVID-19 studyathon. The mapping of data to the OMOP CDM has been supported by a taskforce from the European Health Data and Evidence Network (EHDEN).

## Author Contributions

SFB, MA, TDS mapped source data to the OMOP CDM. EB, CT, and DPA led the data analysis. MR and ER performed a literature review. All authors were involved in the study conception and design, interpretation of the results, and the preparation of the manuscript.

## Ethical approvals

This study was approved by the Clinical Research Ethics Committee of the IDIAPJGol (project code: 20/070-PCV).

## Appendix

**Appendix Figure A1.**
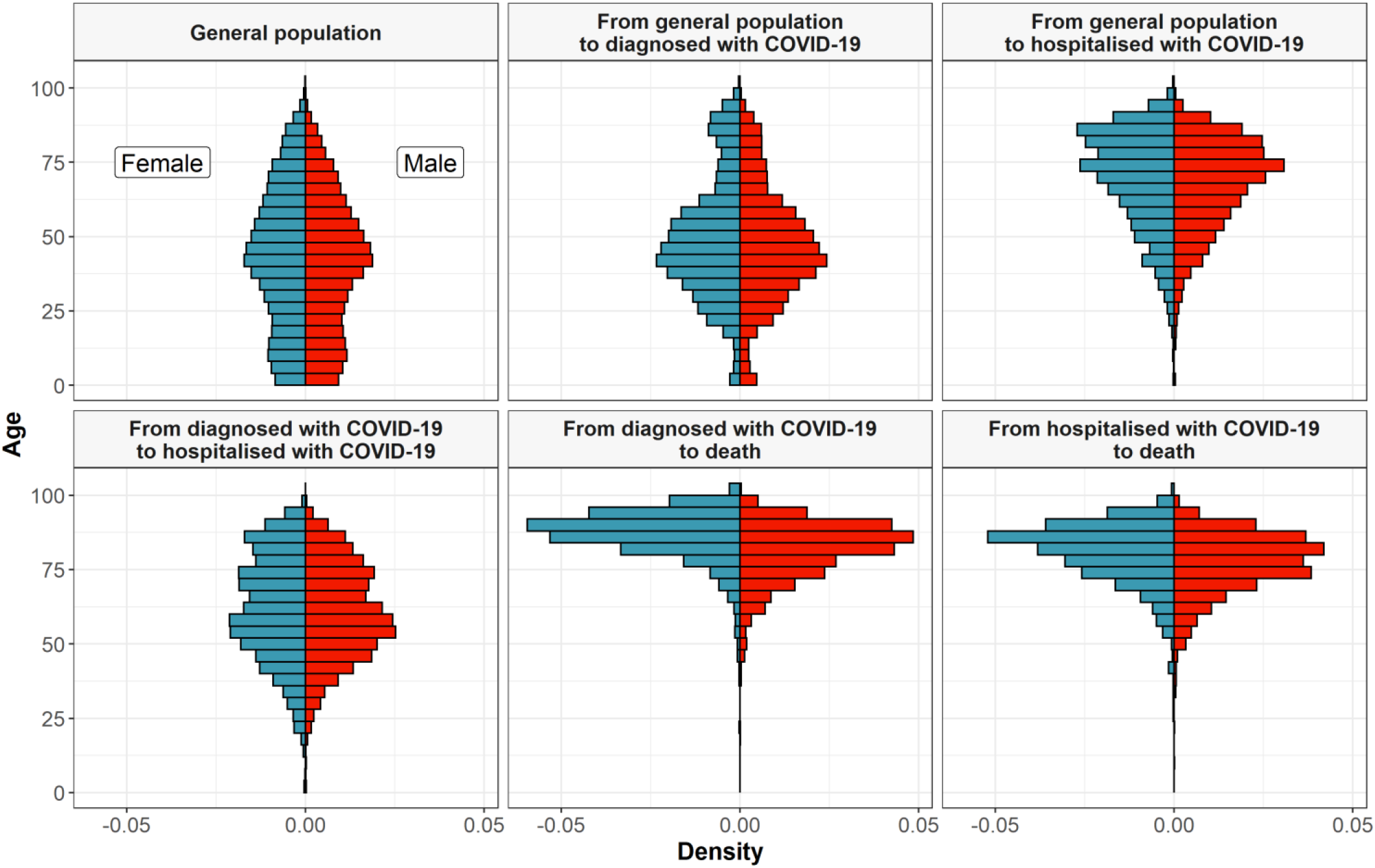
Histogram of age, by gender, for the study population and by transition in the multi-state model

**Appendix Table A1.** Estimated hazard ratios for age: from general population to diagnosed with COVID-19

|  | Overall |  | Male only |  | Female only |  |
| --- | --- | --- | --- | --- | --- | --- |
| Age | Age only model | Age and comorbidity model | Age only model | Age and comorbidity model | Age only model | Age and comorbidity model |
| 20 | 0.69<br>(0.68 to 0.70) | 0.79 (0.77 to 0.81) | 0.65<br>(0.63 to 0.67) | 0.76 (0.74 to 0.79) | 0.74<br>(0.72 to 0.75) | 0.84 (0.82 to 0.86) |
| 25 | 0.93<br>(0.92 to 0.95) | 1.07 (1.04 to 1.09) | 0.84<br>(0.82 to 0.86) | 0.99 (0.95 to 1.02) | 1.03<br>(1.00 to 1.05) | 1.17 (1.14 to 1.20) |
| 30 | 1.21<br>(1.19 to 1.24) | 1.38 (1.35 to 1.41) | 1.06<br>(1.03 to 1.09) | 1.24 (1.20 to 1.28) | 1.37<br>(1.33 to 1.40) | 1.55 (1.51 to 1.59) |
| 35 | 1.49<br>(1.46 to 1.52) | 1.67 (1.64 to 1.71) | 1.28<br>(1.25 to 1.32) | 1.48 (1.44 to 1.53) | 1.70<br>(1.67 to 1.74) | 1.91 (1.86 to 1.96) |
| 40 | 1.66<br>(1.63 to 1.69) | 1.84 (1.81 to 1.88) | 1.42<br>(1.38 to 1.46) | 1.62 (1.57 to 1.68) | 1.92<br>(1.88 to 1.97) | 2.14 (2.09 to 2.20) |
| 45 | 1.62<br>(1.59 to 1.65) | 1.79 (1.75 to 1.82) | 1.39<br>(1.35 to 1.42) | 1.57 (1.52 to 1.61) | 1.90<br>(1.85 to 1.94) | 2.09 (2.04 to 2.15) |
| 50 | 1.39<br>(1.38 to 1.41) | 1.53 (1.50 to 1.55) | 1.22<br>(1.20 to 1.25) | 1.36 (1.33 to 1.39) | 1.61<br>(1.58 to 1.64) | 1.77 (1.73 to 1.80) |
| 55 | 1.15<br>(1.14 to 1.16) | 1.25 (1.23 to 1.26) | 1.06<br>(1.05 to 1.07) | 1.15 (1.14 to 1.17) | 1.28<br>(1.27 to 1.30) | 1.39 (1.37 to 1.40) |
| 60 | 1.02<br>(1.01 to 1.02) | 1.07 (1.06 to 1.07) | 0.99<br>(0.99 to 0.99) | 1.04 (1.03 to 1.04) | 1.06<br>(1.06 to 1.07) | 1.12 (1.11 to 1.12) |
| 65 | ref | ref | ref | ref | ref | ref |
| 70 | 1.08<br>(1.07 to 1.08) | 1.01 (1.01 to 1.01) | 1.07<br>(1.07 to 1.08) | 1.02 (1.01 to 1.03) | 1.06<br>(1.05 to 1.06) | 0.99 (0.98 to 0.99) |
| 75 | 1.23<br>(1.22 to 1.24) | 1.08 (1.07 to 1.09) | 1.20<br>(1.18 to 1.22) | 1.07 (1.05 to 1.09) | 1.22<br>(1.21 to 1.23) | 1.05 (1.03 to 1.06) |
| 80 | 1.47<br>(1.45 to<br>1.49) | 1.18 (1.16 to<br>1.20) | 1.36<br>(1.33 to<br>1.39) | 1.14 (1.11 to<br>1.18) | 1.49<br>(1.46 to<br>1.51) | 1.16 (1.14 to<br>1.18) |
| 85 | 1.77<br>(1.73 to<br>1.80) | 1.31 (1.28 to<br>1.34) | 1.55<br>(1.49 to<br>1.60) | 1.22 (1.17 to<br>1.27) | 1.86<br>(1.81 to<br>1.90) | 1.32 (1.28 to<br>1.36) |
| 90 | 2.13<br>(2.07 to<br>2.18) | 1.45 (1.41 to<br>1.50) | 1.76<br>(1.68 to<br>1.84) | 1.30 (1.24 to<br>1.37) | 2.33<br>(2.25 to<br>2.40) | 1.50 (1.44 to<br>1.55) |
| 95 | 2.56<br>(2.48 to<br>2.64) | 1.61 (1.55 to<br>1.67) | 2.00<br>(1.89 to<br>2.11) | 1.40 (1.31 to<br>1.49) | 2.91<br>(2.80 to<br>3.03) | 1.70 (1.62 to<br>1.78) |

**Appendix Table A2.** Estimated hazard ratios for age: from general population to hospitalised with COVID-19

|  | Overall |  | Male only |  | Female only |  |
| --- | --- | --- | --- | --- | --- | --- |
| Age | Age only model | Age and comorbidity model | Age only model | Age and comorbidity model | Age only model | Age and comorbidity model |
| 20 | 0.037 (0.032 to 0.042) | 0.063 (0.055 to 0.072) | 0.026 (0.022 to 0.032) | 0.043 (0.036 to 0.052) | 0.055 (0.047 to 0.066) | 0.093 (0.078 to 0.11) |
| 25 | 0.058 (0.052 to 0.064) | 0.094 (0.085 to 0.11) | 0.043 (0.037 to 0.05) | 0.068 (0.058 to 0.08) | 0.081 (0.07 to 0.093) | 0.13 (0.11 to 0.15) |
| 30 | 0.089 (0.082 to 0.096) | 0.14 (0.13 to 0.15) | 0.069 (0.062 to 0.078) | 0.10 (0.092 to 0.12) | 0.12 (0.10 to 0.13) | 0.18 (0.16 to 0.20) |
| 35 | 0.13 (0.13 to 0.14) | 0.20 (0.19 to 0.21) | 0.11 (0.10 to 0.12) | 0.16 (0.14 to 0.17) | 0.16 (0.15 to 0.18) | 0.24 (0.22 to 0.27) |
| 40 | 0.20 (0.19 to 0.21) | 0.28 (0.26 to 0.29) | 0.17 (0.16 to 0.18) | 0.23 (0.21 to 0.24) | 0.23 (0.22 to 0.25) | 0.32 (0.30 to 0.34) |
| 45 | 0.28 (0.27 to 0.29) | 0.38 (0.36 to 0.39) | 0.25 (0.24 to 0.26) | 0.32 (0.31 to 0.34) | 0.32 (0.30 to 0.33) | 0.42 (0.40 to 0.44) |
| 50 | 0.40 (0.39 to 0.41) | 0.50 (0.49 to 0.51) | 0.37 (0.36 to 0.38) | 0.44 (0.43 to 0.46) | 0.43 (0.42 to 0.45) | 0.53 (0.52 to 0.55) |
| 55 | 0.56 (0.55 to 0.56) | 0.64 (0.63 to 0.66) | 0.52 (0.51 to 0.53) | 0.60 (0.58 to 0.61) | 0.58 (0.57 to 0.59) | 0.67 (0.65 to 0.69) |
| 60 | 0.75 (0.75 to 0.76) | 0.81 (0.81 to 0.82) | 0.73 (0.73 to 0.74) | 0.78 (0.77 to 0.79) | 0.77 (0.76 to 0.77) | 0.83 (0.82 to 0.84) |
| 65 | ref | ref | ref | ref | ref | ref |
| 70 | 1.30 (1.29 to 1.31) | 1.20 (1.19 to 1.21) | 1.33 (1.32 to 1.35) | 1.24 (1.23 to 1.26) | 1.29 (1.27 to 1.30) | 1.19 (1.17 to 1.20) |
| 75 | 1.65 (1.63 to 1.68) | 1.40 (1.38 to 1.43) | 1.74 (1.70 to 1.78) | 1.51 (1.47 to 1.55) | 1.64 (1.60 to 1.67) | 1.39 (1.35 to 1.43) |
| 80 | 2.06 (2.01 to 2.11) | 1.60 (1.55 to 1.65) | 2.21 (2.13 to 2.30) | 1.78 (1.70 to 1.86) | 2.05 (1.98 to 2.12) | 1.59 (1.52 to 1.67) |
| 85 | 2.51 (2.42 to 2.61) | 1.78 (1.70 to 1.87) | 2.75 (2.61 to 2.91) | 2.05 (1.91 to 2.19) | 2.53 (2.40 to 2.67) | 1.79 (1.67 to 1.92) |
| 90 | 3.00 (2.84 to 3.16) | 1.93 (1.81 to 2.06) | 3.35 (3.10 to 3.61) | 2.29 (2.09 to 2.52) | 3.09 (2.87 to 3.32) | 1.98 (1.80 to 2.17) |
| 95 | 3.51 (3.27 to 3.76) | 2.05 (1.88 to 2.23) | 3.97 (3.58 to 4.40) | 2.51 (2.22 to 2.83) | 3.71 (3.36 to 4.10) | 2.15 (1.90 to 2.43) |

**Appendix Table A3.** Estimated hazard ratios for age: from diagnosed with COVID-19 to hospitalised with COVID-19

|  | Overall |  | Male only |  | Female only |  |
| --- | --- | --- | --- | --- | --- | --- |
| Age | Age only model | Age and comorbidity model | Age only model | Age and comorbidity model | Age only model | Age and comorbidity model |
| 20 | 0.076<br>(0.067 to 0.086) | 0.092 (0.08 to 0.10) | 0.056<br>(0.046 to 0.067) | 0.063 (0.051 to 0.076) | 0.10<br>(0.084 to 0.12) | 0.12 (0.10 to 0.15) |
| 25 | 0.099<br>(0.091 to 0.11) | 0.12 (0.11 to 0.13) | 0.078<br>(0.068 to 0.089) | 0.087 (0.076 to 0.10) | 0.12<br>(0.11 to 0.14) | 0.15 (0.13 to 0.17) |
| 30 | 0.13 (0.12 to 0.14) | 0.15 (0.14 to 0.16) | 0.11<br>(0.097 to 0.12) | 0.12 (0.11 to 0.13) | 0.15<br>(0.14 to 0.17) | 0.18 (0.16 to 0.20) |
| 35 | 0.16 (0.15 to 0.17) | 0.19 (0.18 to 0.20) | 0.15 (0.13 to 0.16) | 0.16 (0.15 to 0.18) | 0.18<br>(0.16 to 0.20) | 0.21 (0.19 to 0.23) |
| 40 | 0.19 (0.18 to 0.20) | 0.23 (0.21 to 0.24) | 0.20 (0.18 to 0.21) | 0.22 (0.20 to 0.24) | 0.20<br>(0.18 to 0.22) | 0.23 (0.21 to 0.25) |
| 45 | 0.24 (0.22 to 0.25) | 0.28 (0.26 to 0.30) | 0.27 (0.25 to 0.29) | 0.30 (0.27 to 0.33) | 0.22<br>(0.20 to 0.24) | 0.26 (0.24 to 0.29) |
| 50 | 0.34 (0.32 to 0.36) | 0.39 (0.36 to 0.41) | 0.40 (0.37 to 0.43) | 0.43 (0.40 to 0.47) | 0.29<br>(0.27 to 0.32) | 0.34 (0.31 to 0.37) |
| 55 | 0.53 (0.51 to 0.55) | 0.58 (0.55 to 0.60) | 0.60 (0.57 to 0.63) | 0.62 (0.59 to 0.65) | 0.47<br>(0.44 to 0.49) | 0.52 (0.48 to 0.55) |
| 60 | 0.79 (0.78 to 0.80) | 0.81 (0.80 to 0.83) | 0.83 (0.81 to 0.84) | 0.84 (0.82 to 0.85) | 0.74<br>(0.72 to 0.76) | 0.77 (0.75 to 0.79) |
| 65 | ref | ref | ref | ref | ref | ref |
| 70 | 1.10 (1.09 to 1.11) | 1.09 (1.08 to 1.10) | 1.08 (1.07 to 1.10) | 1.09 (1.07 to 1.11) | 1.14<br>(1.13 to 1.16) | 1.13 (1.11 to 1.15) |
| 75 | 1.07 (1.05 to 1.09) | 1.07 (1.05 to 1.10) | 1.08 (1.05 to 1.11) | 1.11 (1.07 to 1.15) | 1.14<br>(1.11 to 1.17) | 1.13 (1.09 to 1.17) |
| 80 | 0.97 (0.94 to 1.00) | 0.99 (0.95 to 1.03) | 1.03 (0.98 to 1.08) | 1.08 (1.02 to 1.15) | 1.02<br>(0.98 to 1.07) | 1.04 (0.99 to 1.10) |
| 85 | 0.83 (0.79 to 0.87) | 0.87 (0.82 to 0.92) | 0.96 (0.89 to 1.03) | 1.03 (0.95 to 1.13) | 0.86 (0.81 to 0.92) | 0.91 (0.84 to 0.98) |
| 90 | 0.70 (0.65 to 0.74) | 0.75 (0.70 to 0.82) | 0.89 (0.80 to 0.98) | 0.98 (0.88 to 1.10) | 0.70 (0.64 to 0.77) | 0.77 (0.69 to 0.86) |
| 95 | 0.58 (0.54 to 0.64) | 0.65 (0.59 to 0.72) | 0.82 (0.72 to 0.93) | 0.94 (0.81 to 1.08) | 0.57 (0.50 to 0.64) | 0.65 (0.56 to 0.75) |

**Appendix Table A4.**
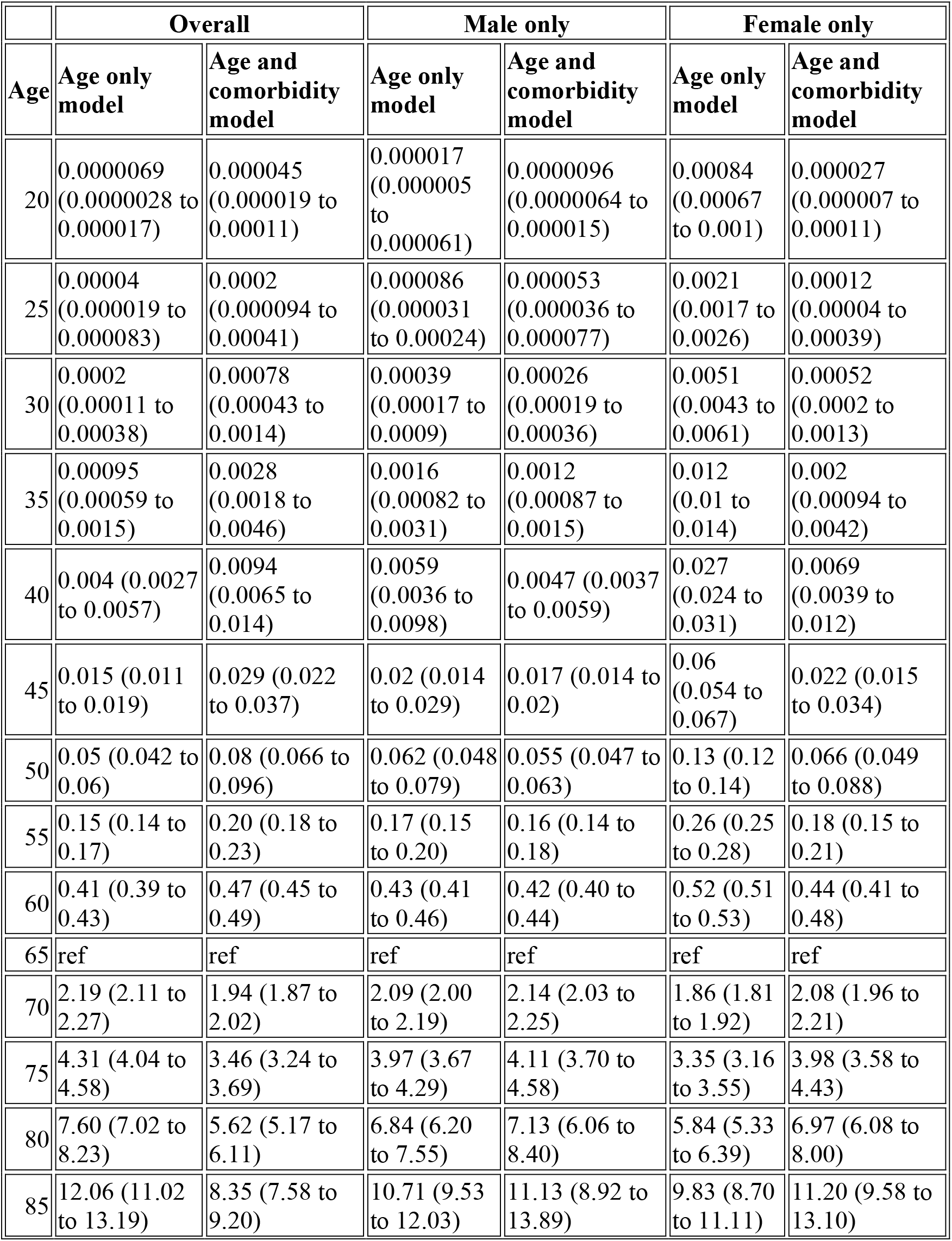
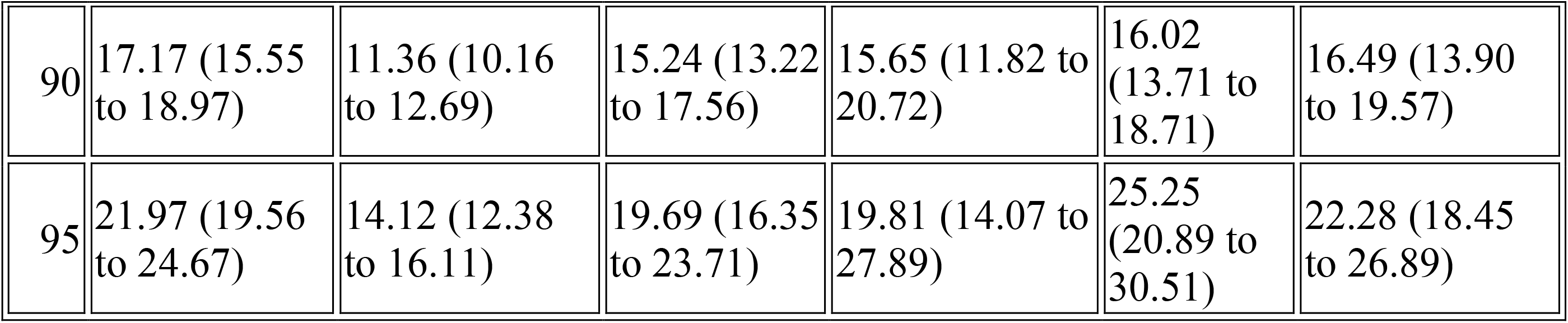
Estimated hazard ratios for age: from diagnosed with COVID-19 to death

|  | Overall |  | Male only |  | Female only |  |
| --- | --- | --- | --- | --- | --- | --- |
| Age | Age only model | Age and comorbidity model | Age only model | Age and comorbidity model | Age only model | Age and comorbidity model |
| 20 | 0.0000069<br>(0.0000028 to 0.000017) | 0.000045<br>(0.000019 to 0.00011) | 0.000017<br>(0.000005 to 0.000061) | 0.0000096<br>(0.0000064 to 0.000015) | 0.00084<br>(0.00067 to 0.001) | 0.000027<br>(0.000007 to 0.00011) |
| 25 | 0.00004<br>(0.000019 to 0.000083) | 0.0002<br>(0.000094 to 0.00041) | 0.000086<br>(0.000031 to 0.00024) | 0.000053<br>(0.000036 to 0.000077) | 0.0021<br>(0.0017 to 0.0026) | 0.00012<br>(0.00004 to 0.00039) |
| 30 | 0.0002<br>(0.00011 to 0.00038) | 0.00078<br>(0.00043 to 0.0014) | 0.00039<br>(0.00017 to 0.0009) | 0.00026<br>(0.00019 to 0.00036) | 0.0051<br>(0.0043 to 0.0061) | 0.00052<br>(0.0002 to 0.0013) |
| 35 | 0.00095<br>(0.00059 to 0.0015) | 0.0028<br>(0.0018 to 0.0046) | 0.0016<br>(0.00082 to 0.0031) | 0.0012<br>(0.00087 to 0.0015) | 0.012<br>(0.01 to 0.014) | 0.002<br>(0.00094 to 0.0042) |
| 40 | 0.004 (0.0027 to 0.0057) | 0.0094<br>(0.0065 to 0.014) | 0.0059<br>(0.0036 to 0.0098) | 0.0047 (0.0037 to 0.0059) | 0.027<br>(0.024 to 0.031) | 0.0069<br>(0.0039 to 0.012) |
| 45 | 0.015 (0.011 to 0.019) | 0.029 (0.022 to 0.037) | 0.02 (0.014 to 0.029) | 0.017 (0.014 to 0.02) | 0.06<br>(0.054 to 0.067) | 0.022 (0.015 to 0.034) |
| 50 | 0.05 (0.042 to 0.06) | 0.08 (0.066 to 0.096) | 0.062 (0.048 to 0.079) | 0.055 (0.047 to 0.063) | 0.13 (0.12 to 0.14) | 0.066 (0.049 to 0.088) |
| 55 | 0.15 (0.14 to 0.17) | 0.20 (0.18 to 0.23) | 0.17 (0.15 to 0.20) | 0.16 (0.14 to 0.18) | 0.26 (0.25 to 0.28) | 0.18 (0.15 to 0.21) |
| 60 | 0.41 (0.39 to 0.43) | 0.47 (0.45 to 0.49) | 0.43 (0.41 to 0.46) | 0.42 (0.40 to 0.44) | 0.52 (0.51 to 0.53) | 0.44 (0.41 to 0.48) |
| 65 | ref | ref | ref | ref | ref | ref |
| 70 | 2.19 (2.11 to 2.27) | 1.94 (1.87 to 2.02) | 2.09 (2.00 to 2.19) | 2.14 (2.03 to 2.25) | 1.86 (1.81 to 1.92) | 2.08 (1.96 to 2.21) |
| 75 | 4.31 (4.04 to 4.58) | 3.46 (3.24 to 3.69) | 3.97 (3.67 to 4.29) | 4.11 (3.70 to 4.58) | 3.35 (3.16 to 3.55) | 3.98 (3.58 to 4.43) |
| 80 | 7.60 (7.02 to 8.23) | 5.62 (5.17 to 6.11) | 6.84 (6.20 to 7.55) | 7.13 (6.06 to 8.40) | 5.84 (5.33 to 6.39) | 6.97 (6.08 to 8.00) |
| 85 | 12.06 (11.02 to 13.19) | 8.35 (7.58 to 9.20) | 10.71 (9.53 to 12.03) | 11.13 (8.92 to 13.89) | 9.83 (8.70 to 11.11) | 11.20 (9.58 to 13.10) |
| 90 | 17.17 (15.55 to 18.97) | 11.36 (10.16 to 12.69) | 15.24 (13.22 to 17.56) | 15.65 (11.82 to 20.72) | 16.02 (13.71 to 18.71) | 16.49 (13.90 to 19.57) |
| 95 | 21.97 (19.56 to 24.67) | 14.12 (12.38 to 16.11) | 19.69 (16.35 to 23.71) | 19.81 (14.07 to 27.89) | 25.25 (20.89 to 30.51) | 22.28 (18.45 to 26.89) |

**Appendix Table A5.** Estimated hazard ratios for age: from hospitalised with COVID-19 to death

|  | Overall |  | Male |  | Female |  |
| --- | --- | --- | --- | --- | --- | --- |
| Age | Age only model | Age and comorbidity model | Age only model | Age and comorbidity model | Age only model | Age and comorbidity model |
| 20 | 0.005<br>(0.0025 to 0.01) | 0.012 (0.0062 to 0.024) | 0.0063<br>(0.0026 to 0.015) | 0.014 (0.0056 to 0.033) | 0.0042<br>(0.0013 to 0.014) | 0.0092 (0.0029 to 0.029) |
| 25 | 0.01<br>(0.0058 to 0.018) | 0.022 (0.013 to 0.039) | 0.012<br>(0.006 to 0.025) | 0.024 (0.012 to 0.05) | 0.0087<br>(0.0033 to 0.023) | 0.017 (0.0066 to 0.045) |
| 30 | 0.02<br>(0.013 to 0.032) | 0.039 (0.025 to 0.062) | 0.024<br>(0.013 to 0.042) | 0.042 (0.023 to 0.075) | 0.017<br>(0.008 to 0.038) | 0.031 (0.014 to 0.068) |
| 35 | 0.039<br>(0.027 to 0.056) | 0.067 (0.047 to 0.096) | 0.044<br>(0.028 to 0.069) | 0.07 (0.044 to 0.11) | 0.034<br>(0.018 to 0.063) | 0.056 (0.03 to 0.10) |
| 40 | 0.072<br>(0.055 to 0.095) | 0.11 (0.086 to 0.15) | 0.079<br>(0.056 to 0.11) | 0.12 (0.082 to 0.16) | 0.064<br>(0.04 to 0.10) | 0.096 (0.061 to 0.15) |
| 45 | 0.13 (0.11 to 0.16) | 0.18 (0.15 to 0.22) | 0.14 (0.11 to 0.18) | 0.19 (0.15 to 0.24) | 0.12<br>(0.084 to 0.17) | 0.16 (0.12 to 0.23) |
| 50 | 0.23 (0.20 to 0.26) | 0.29 (0.25 to 0.33) | 0.24 (0.20 to 0.28) | 0.29 (0.25 to 0.35) | 0.21 (0.17 to 0.26) | 0.27 (0.21 to 0.33) |
| 55 | 0.38 (0.35 to 0.41) | 0.45 (0.42 to 0.49) | 0.39 (0.36 to 0.43) | 0.45 (0.41 to 0.50) | 0.36 (0.32 to 0.42) | 0.42 (0.37 to 0.49) |
| 60 | 0.63 (0.61 to 0.65) | 0.68 (0.66 to 0.70) | 0.64 (0.61 to 0.66) | 0.68 (0.65 to 0.71) | 0.61 (0.58 to 0.65) | 0.66 (0.62 to 0.70) |
| 65 | ref | ref | ref | ref | ref | ref |
| 70 | 1.54 (1.51 to 1.58) | 1.43 (1.40 to 1.47) | 1.53 (1.48 to 1.57) | 1.44 (1.39 to 1.48) | 1.59 (1.52 to 1.66) | 1.48 (1.41 to 1.54) |
| 75 | 2.31 (2.21 to 2.41) | 2.00 (1.92 to 2.10) | 2.27 (2.16 to 2.39) | 2.02 (1.91 to 2.14) | 2.45 (2.27 to 2.64) | 2.12 (1.96 to 2.29) |
| 80 | 3.35 (3.17 to 3.53) | 2.73 (2.57 to 2.91) | 3.29 (3.07 to 3.52) | 2.77 (2.55 to 3.00) | 3.67 (3.33 to 4.04) | 2.97 (2.67 to 3.29) |
| 85 | 4.70 (4.38 to 5.05) | 3.63 (3.34 to 3.93) | 4.62 (4.21 to 5.07) | 3.71 (3.33 to 4.14) | 5.34 (4.76 to 5.99) | 4.04 (3.54 to 4.61) |
| 90 | 6.41 (5.84 to 7.04) | 4.69 (4.21 to 5.22) | 6.32 (5.54 to 7.19) | 4.86 (4.19 to 5.64) | 7.55 (6.55 to 8.70) | 5.36 (4.54 to 6.33) |
| 95 | 8.47 (7.46 to 9.62) | 5.92 (5.12 to 6.84) | 8.40 (7.00 to 10.08) | 6.23 (5.08 to 7.63) | 10.37 (8.61 to 12.48) | 6.93 (5.59 to 8.60) |

**Appendix Figure A2.**
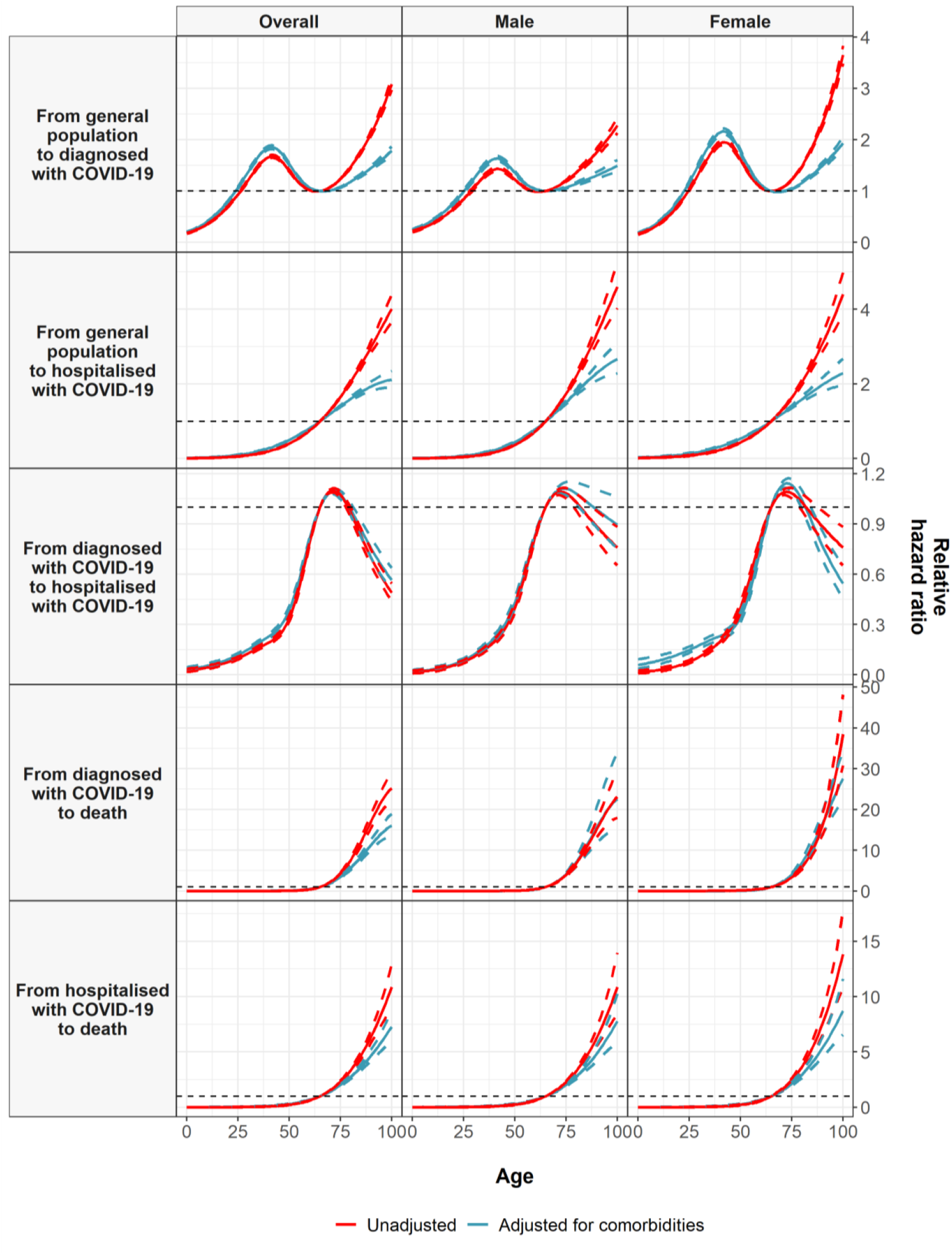
Estimated hazard ratios for age, with a reference age of 65.

**Appendix Table A5.**
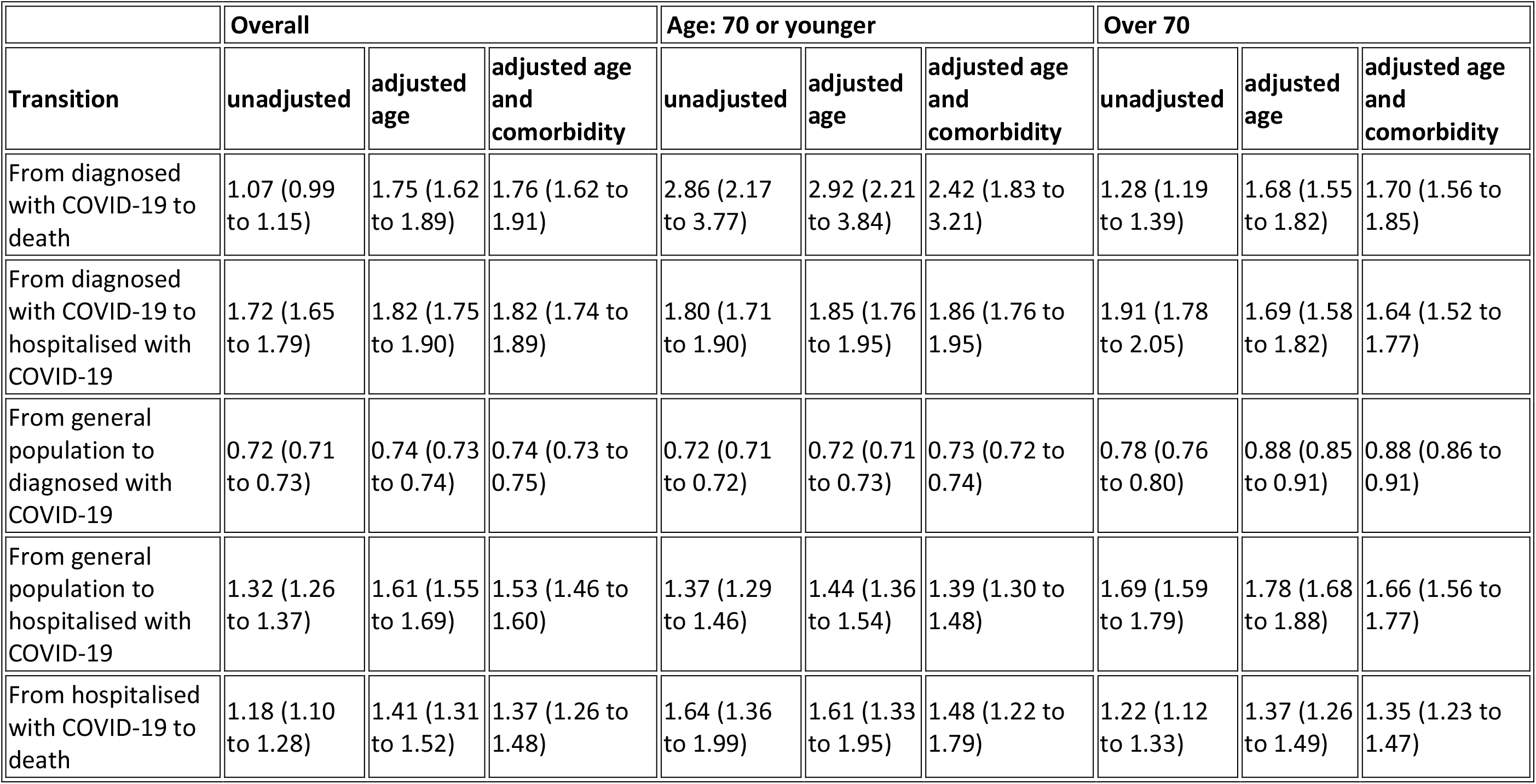
Estimated hazard ratios for male gender

|  | Overall |  |  | Age: 70 or younger |  |  | Over 70 |  |  |
| --- | --- | --- | --- | --- | --- | --- | --- | --- | --- |
| Transition | unadjusted | adjusted age | adjusted age and comorbidity | unadjusted | adjusted age | adjusted age and comorbidity | unadjusted | adjusted age | adjusted age and comorbidity |
| From diagnosed with COVID-19 to death | 1.07 (0.99 to 1.15) | 1.75 (1.62 to 1.89) | 1.76 (1.62 to 1.91) | 2.86 (2.17 to 3.77) | 2.92 (2.21 to 3.84) | 2.42 (1.83 to 3.21) | 1.28 (1.19 to 1.39) | 1.68 (1.55 to 1.82) | 1.70 (1.56 to 1.85) |
| From diagnosed with COVID-19 to hospitalised with COVID-19 | 1.72 (1.65 to 1.79) | 1.82 (1.75 to 1.90) | 1.82 (1.74 to 1.89) | 1.80 (1.71 to 1.90) | 1.85 (1.76 to 1.95) | 1.86 (1.76 to 1.95) | 1.91 (1.78 to 2.05) | 1.69 (1.58 to 1.82) | 1.64 (1.52 to 1.77) |
| From general population to diagnosed with COVID-19 | 0.72 (0.71 to 0.73) | 0.74 (0.73 to 0.74) | 0.74 (0.73 to 0.75) | 0.72 (0.71 to 0.72) | 0.72 (0.71 to 0.73) | 0.73 (0.72 to 0.74) | 0.78 (0.76 to 0.80) | 0.88 (0.85 to 0.91) | 0.88 (0.86 to 0.91) |
| From general population to hospitalised with COVID-19 | 1.32 (1.26 to 1.37) | 1.61 (1.55 to 1.69) | 1.53 (1.46 to 1.60) | 1.37 (1.29 to 1.46) | 1.44 (1.36 to 1.54) | 1.39 (1.30 to 1.48) | 1.69 (1.59 to 1.79) | 1.78 (1.68 to 1.88) | 1.66 (1.56 to 1.77) |
| From hospitalised with COVID-19 to death | 1.18 (1.10 to 1.28) | 1.41 (1.31 to 1.52) | 1.37 (1.26 to 1.48) | 1.64 (1.36 to 1.99) | 1.61 (1.33 to 1.95) | 1.48 (1.22 to 1.79) | 1.22 (1.12 to 1.33) | 1.37 (1.26 to 1.49) | 1.35 (1.23 to 1.47) |

**Appendix Figure A3.**
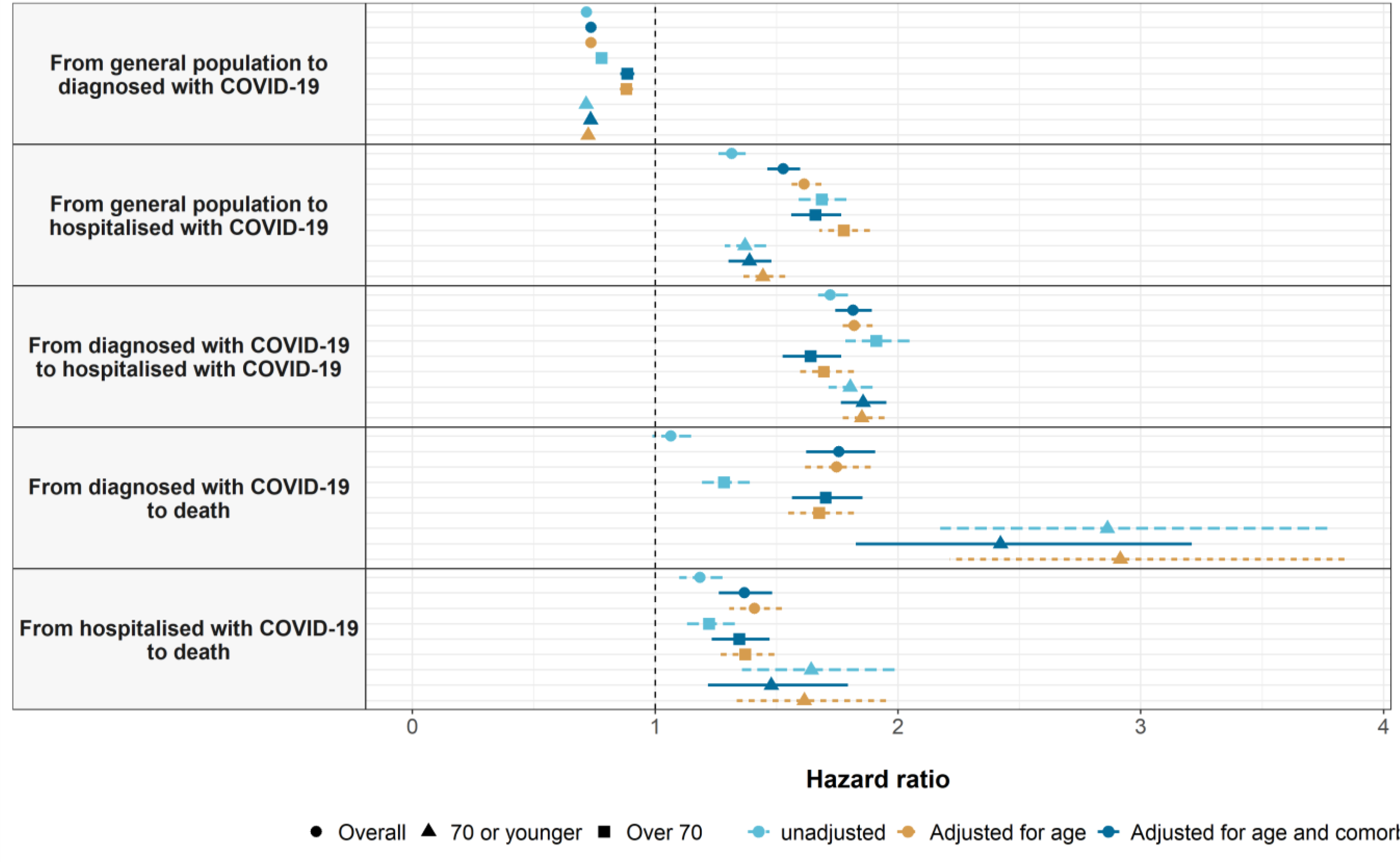
Estimated hazard ratios for male, relative to female, gender

**Appendix Table A6.** Estimated hazard ratios for comorbidities

|  |  | Overall |  | Male- 70 or younger |  | Female- 70 or younger |  | Male- Over 70 |  | Female- Over 70 |  |
| --- | --- | --- | --- | --- | --- | --- | --- | --- | --- | --- | --- |
| Transition | Variable | Unadjusted | Adjusted for age and gender | Unadjusted | Adjusted for age | Unadjusted | Adjusted for age | Unadjusted | Adjusted for age | Unadjusted | Adjusted for age |
| From general population to diagnosed with COVID-19 | Charlson - 1:0 | 1.47 (1.44 to 1.50) | 1.35 (1.32 to 1.38) | 1.31 (1.26 to 1.36) | 1.07 (1.03 to 1.11) | 1.41 (1.36 to 1.45) | 1.19 (1.16 to 1.23) | 1.68 (1.55 to 1.82) | 1.58 (1.45 to 1.71) | 2.53 (2.39 to 2.67) | 2.11 (2.00 to 2.23) |
|  | Charlson - 2:0 | 1.27 (1.24 to 1.30) | 1.13 (1.10 to 1.16) | 1.25 (1.19 to 1.31) | 0.92 (0.87 to 0.97) | 1.21 (1.16 to 1.25) | 0.92 (0.88 to 0.95) | 1.54 (1.43 to 1.66) | 1.39 (1.29 to 1.50) | 1.71 (1.62 to 1.80) | 1.42 (1.34 to 1.50) |
|  | Charlson - 3+:0 | 1.74 (1.71 to 1.78) | 1.51 (1.47 to 1.55) | 1.29 (1.22 to 1.37) | 0.93 (0.87 to 0.98) | 1.31 (1.23 to 1.40) | 0.96 (0.90 to 1.03) | 2.13 (2.00 to 2.28) | 1.78 (1.66 to 1.91) | 2.91 (2.77 to 3.06) | 2.06 (1.96 to 2.17) |
|  | Autoimmune condition | 1.31 (1.28 to 1.35) | 1.14 (1.11 to 1.17) | 1.31 (1.25 to 1.37) | 1.14 (1.09 to 1.20) | 1.33 (1.28 to 1.38) | 1.17 (1.13 to 1.21) | 1.10 (1.01 to 1.19) | 1.08 (0.99 to 1.18) | 1.08 (1.02 to 1.14) | 1.07 (1.01 to 1.13) |
|  | Chronic kidney disease | 1.46 (1.42 to 1.50) | 1.08 (1.05 to 1.12) | 1.21 (1.11 to 1.32) | 0.87 (0.79 to 0.95) | 1.06 (0.97 to 1.17) | 0.78 (0.71 to 0.85) | 1.25 (1.19 to 1.33) | 1.06 (1.00 to 1.12) | 1.42 (1.36 to 1.48) | 1.03 (0.99 to 1.07) |
|  | COPD | 1.25 (1.21 to 1.30) | 1.16 (1.12 to 1.21) | 1.12 (1.04 to 1.21) | 0.80 (0.74 to 0.86) | 1.20 (1.10 to 1.30) | 0.88 (0.81 to 0.96) | 1.29 (1.21 to 1.38) | 1.26 (1.18 to 1.35) | 1.27 (1.17 to 1.39) | 1.21 (1.11 to 1.32) |
|  | Dementia | 4.72 (4.58 to 4.85) | 3.65 (3.53 to 3.77) | NA | NA | NA | NA | 4.24 (3.97 to 4.52) | 3.48 (3.26 to 3.72) | 4.91 (4.71 to 5.10) | 3.55 (3.40 to 3.70) |
|  | Heart disease | 1.32 (1.30 to 1.34) | 1.14 (1.12 to 1.17) | 1.25 (1.20 to 1.30) | 0.98 (0.94 to 1.02) | 1.33 (1.28 to 1.38) | 1.05 (1.01 to 1.09) | 1.23 (1.17 to 1.29) | 1.09 (1.03 to 1.14) | 1.30 (1.25 to 1.34) | 1.05 (1.02 to 1.09) |
|  | Hyperlipidemia | 1.22 (1.19 to 1.24) | 1.09 (1.07 to 1.11) | 1.37 (1.32 to 1.41) | 1.09 (1.06 to 1.13) | 1.22 (1.18 to 1.26) | 0.94 (0.91 to 0.97) | 0.96 (0.90 to 1.03) | 1.05 (0.99 to 1.13) | 0.84 (0.80 to 0.88) | 0.96 (0.92 to 1.01) |
|  | Hypertension | 1.24 (1.22 to 1.26) | 1.10 (1.08 to 1.12) | 1.32 (1.28 to 1.36) | 0.97 (0.94 to 1.00) | 1.23 (1.20 to 1.27) | 0.91 (0.89 to 0.94) | 0.99 (0.94 to 1.04) | 1.02 (0.97 to 1.08) | 1.08 (1.04 to 1.12) | 1.08 (1.04 to 1.12) |
|  | Malignant neoplasm | 1.18 (1.16 to 1.21) | 0.99 (0.97 to 1.02) | 1.18 (1.11 to 1.25) | 0.86 (0.81 to 0.91) | 1.13 (1.08 to 1.18) | 0.87 (0.83 to 0.91) | 1.05 (1.00 to 1.11) | 0.99 (0.94 to 1.05) | 0.99 (0.94 to 1.04) | 0.95 (0.90 to 0.99) |
|  | Obesity | 1.30 (1.28 to 1.32) | 1.17 (1.15 to 1.18) | 1.35 (1.32 to 1.39) | 1.14 (1.11 to 1.17) | 1.41 (1.38 to 1.44) | 1.22 (1.19 to 1.24) | 0.91 (0.86 to 0.96) | 0.97 (0.92 to 1.02) | 0.83 (0.80 to 0.86) | 0.94 (0.91 to 0.98) |
|  | Type 2 diabetes | 1.21 (1.19 to 1.24) | 1.08 (1.06 to 1.11) | 1.21 (1.15 to 1.26) | 0.86 (0.82 to 0.90) | 1.20 (1.14 to 1.26) | 0.88 (0.83 to 0.92) | 1.08 (1.02 to 1.14) | 1.12 (1.06 to 1.19) | 1.11 (1.06 to 1.16) | 1.13 (1.08 to 1.19) |
| From general population to hospitalised with COVID-19 | Charlson - 1:0 | 3.46 (3.24 to 3.69) | 1.49 (1.39 to 1.60) | 2.62 (2.31 to 2.97) | 1.29 (1.14 to 1.47) | 2.30 (1.96 to 2.70) | 1.42 (1.21 to 1.67) | 1.57 (1.37 to 1.79) | 1.53 (1.33 to 1.75) | 1.65 (1.44 to 1.89) | 1.55 (1.35 to 1.78) |
|  | Charlson - 2:0 | 5.13 (4.83 to 5.44) | 1.64 (1.54 to 1.75) | 4.44 (3.91 to 5.03) | 1.61 (1.41 to 1.83) | 3.92 (3.40 to 4.52) | 1.81 (1.57 to 2.10) | 1.56 (1.38 to 1.76) | 1.50 (1.32 to 1.69) | 1.66 (1.47 to 1.88) | 1.56 (1.38 to 1.76) |
|  | Charlson - 3+:0 | 9.46 (8.97 to 9.97) | 2.31 (2.17 to 2.46) | 6.17 (5.40 to 7.04) | 2.05 (1.79 to 2.36) | 7.30 (6.09 to 8.75) | 3.04 (2.52 to 3.66) | 2.27 (2.04 to 2.53) | 2.11 (1.89 to 2.36) | 2.57 (2.30 to 2.87) | 2.28 (2.03 to 2.56) |
|  | Autoimmune condition | 2.19 (2.04 to 2.35) | 1.33 (1.24 to 1.43) | 2.11 (1.81 to 2.46) | 1.34 (1.15 to 1.56) | 2.33 (1.99 to 2.72) | 1.60 (1.36 to 1.87) | 1.24 (1.09 to 1.42) | 1.23 (1.08 to 1.41) | 1.29 (1.13 to 1.46) | 1.28 (1.12 to 1.45) |
|  | Chronic kidney disease | 5.68 (5.37 to 6.00) | 1.47 (1.38 to 1.56) | 5.87 (4.96 to 6.94) | 1.99 (1.68 to 2.37) | 6.02 (4.80 to 7.55) | 2.39 (1.90 to 3.01) | 1.45 (1.33 to 1.58) | 1.34 (1.22 to 1.46) | 1.57 (1.43 to 1.73) | 1.37 (1.25 to 1.51) |
|  | COPD | 4.69 (4.36 to 5.04) | 1.51 (1.40 to 1.63) | 4.07 (3.45 to 4.80) | 1.37 (1.16 to 1.62) | 3.49 (2.62 to 4.64) | 1.56 (1.17 to 2.07) | 1.45 (1.31 to 1.60) | 1.43 (1.29 to 1.58) | 1.68 (1.41 to 2.00) | 1.65 (1.38 to 1.97) |
|  | Dementia | 7.48 (6.89 to 8.13) | 1.82 (1.67 to 1.99) | NA | NA | NA | NA | 1.92 (1.67 to 2.21) | 1.71 (1.49 to 1.97) | 2.23 (1.99 to 2.51) | 1.91 (1.69 to 2.15) |
|  | Heart disease | 4.63 (4.43 to 4.84) | 1.42 (1.35 to 1.49) | 3.24 (2.92 to 3.60) | 1.35 (1.21 to 1.51) | 3.07 (2.64 to 3.57) | 1.57 (1.35 to 1.83) | 1.43 (1.32 to 1.54) | 1.35 (1.24 to 1.46) | 1.53 (1.41 to 1.67) | 1.40 (1.28 to 1.53) |
|  | Hyperlipidemia | 2.24 (2.12 to 2.36) | 1.16 (1.10 to 1.23) | 2.77 (2.50 to 3.06) | 1.33 (1.20 to 1.47) | 2.62 (2.31 to 2.97) | 1.21 (1.06 to 1.38) | 1.11 (1.00 to 1.23) | 1.16 (1.05 to 1.28) | 0.92 (0.82 to 1.03) | 0.97 (0.87 to 1.09) |
|  | Hypertension | 3.07 (2.93 to 3.21) | 1.11 (1.06 to 1.17) | 3.19 (2.91 to 3.50) | 1.16 (1.05 to 1.27) | 3.41 (3.05 to 3.83) | 1.49 (1.32 to 1.67) | 0.98 (0.90 to 1.06) | 0.99 (0.92 to 1.08) | 1.01 (0.93 to 1.11) | 1.01 (0.92 to 1.11) |
|  | Malignant neoplasm | 3.53 (3.34 to 3.74) | 1.21 (1.14 to 1.28) | 3.63 (3.16 to 4.18) | 1.31 (1.13 to 1.51) | 3.05 (2.60 to 3.57) | 1.45 (1.23 to 1.71) | 1.13 (1.03 to 1.23) | 1.09 (1.00 to 1.19) | 1.16 (1.03 to 1.30) | 1.13 (1.01 to 1.27) |
|  | Obesity | 3.27 (3.13 to 3.41) | 1.74 (1.66 to 1.82) | 3.54 (3.25 to 3.86) | 1.85 (1.69 to 2.02) | 4.18 (3.80 to 4.61) | 2.67 (2.42 to 2.94) | 1.28 (1.18 to 1.39) | 1.33 (1.22 to 1.44) | 1.49 (1.36 to 1.62) | 1.57 (1.44 to 1.71) |
|  | Type 2 diabetes | 4.28 (4.06 to 4.51) | 1.53 (1.45 to 1.62) | 4.49 (4.04 to 5.00) | 1.63 (1.46 to 1.82) | 5.61 (4.88 to 6.44) | 2.43 (2.11 to 2.81) | 1.30 (1.19 to 1.42) | 1.32 (1.21 to 1.44) | 1.34 (1.21 to 1.49) | 1.35 (1.21 to 1.50) |
| From diagnosed with COVID-19 to hospitalised with COVID-19 | Charlson - 1:0 | 1.67 (1.56 to 1.78) | 0.99 (0.93 to 1.06) | 1.53 (1.37 to 1.70) | 0.98 (0.88 to 1.09) | 1.44 (1.27 to 1.63) | 1.11 (0.98 to 1.26) | 0.80 (0.68 to 0.95) | 0.88 (0.74 to 1.04) | 0.73 (0.63 to 0.85) | 0.88 (0.76 to 1.02) |
|  | Charlson - 2:0 | 2.44 (2.30 to 2.60) | 1.16 (1.09 to 1.24) | 2.44 (2.17 to 2.75) | 1.15 (1.01 to 1.29) | 1.90 (1.66 to 2.18) | 1.16 (1.01 to 1.33) | 1.03 (0.89 to 1.20) | 1.10 (0.95 to 1.28) | 0.88 (0.77 to 1.01) | 1.03 (0.90 to 1.19) |
|  | Charlson - 3+:0 | 2.86 (2.70 to 3.03) | 1.19 (1.11 to 1.28) | 2.40 (2.08 to 2.77) | 1.01 (0.87 to 1.17) | 3.07 (2.55 to 3.69) | 1.60 (1.33 to 1.93) | 0.95 (0.83 to 1.08) | 1.09 (0.95 to 1.24) | 0.83 (0.74 to 0.95) | 1.11 (0.98 to 1.27) |
|  | Autoimmune condition | 1.37 (1.27 to 1.47) | 1.08 (1.01 to 1.17) | 1.51 (1.33 to 1.72) | 1.19 (1.05 to 1.36) | 1.24 (1.07 to 1.43) | 1.01 (0.87 to 1.17) | 0.94 (0.79 to 1.12) | 0.93 (0.78 to 1.11) | 1.17 (1.01 to 1.36) | 1.13 (0.98 to 1.31) |
|  | Chronic kidney disease | 2.45 (2.30 to 2.62) | 1.24 (1.16 to 1.34) | 6.48 (5.69 to 7.37) | 1.28 (1.06 to 1.54) | 3.50 (2.77 to 4.42) | 1.84 (1.45 to 2.33) | 1.09 (0.97 to 1.22) | 1.18 (1.05 to 1.32) | 0.96 (0.86 to 1.07) | 1.18 (1.05 to 1.32) |
|  | COPD | 2.34 (2.14 to 2.56) | 1.00 (0.91 to 1.10) | 1.99 (1.66 to 2.39) | 0.88 (0.73 to 1.06) | 1.43 (1.03 to 1.99) | 0.76 (0.54 to 1.07) | 1.03 (0.90 to 1.17) | 1.03 (0.91 to 1.18) | 1.11 (0.89 to 1.38) | 1.11 (0.89 to 1.38) |
|  | Dementia | 1.27 (1.16 to 1.40) | 0.59 (0.54 to 0.65) | NA | NA | NA | NA | 0.48 (0.40 to 0.56) | 0.52 (0.44 to 0.62) | 0.52 (0.46 to 0.59) | 0.63 (0.55 to 0.71) |
|  | Heart disease | 2.21 (2.10 to 2.31) | 1.10 (1.05 to 1.16) | 1.78 (1.61 to 1.97) | 0.98 (0.89 to 1.09) | 1.65 (1.44 to 1.90) | 1.07 (0.93 to 1.24) | 1.05 (0.95 to 1.15) | 1.12 (1.01 to 1.24) | 1.04 (0.95 to 1.15) | 1.16 (1.05 to 1.28) |
|  | Hyperlipidemia | 1.73 (1.64 to 1.82) | 1.09 (1.04 to 1.16) | 1.72 (1.58 to 1.88) | 1.12 (1.02 to 1.22) | 1.91 (1.72 to 2.12) | 1.17 (1.05 to 1.30) | 1.16 (1.02 to 1.31) | 1.09 (0.96 to 1.24) | 1.09 (0.96 to 1.24) | 0.99 (0.87 to 1.13) |
|  | Hypertension | 2.04 (1.95 to 2.14) | 1.05 (1.00 to 1.10) | 2.10 (1.94 to 2.27) | 1.07 (0.98 to 1.16) | 2.17 (1.97 to 2.39) | 1.30 (1.17 to 1.43) | 0.91 (0.82 to 1.01) | 0.90 (0.82 to 1.00) | 0.96 (0.87 to 1.06) | 0.98 (0.89 to 1.09) |
|  | Malignant neoplasm | 2.10 (1.97 to 2.24) | 1.08 (1.01 to 1.15) | 2.01 (1.75 to 2.32) | 0.93 (0.81 to 1.08) | 1.86 (1.61 to 2.16) | 1.18 (1.01 to 1.37) | 1.05 (0.94 to 1.18) | 1.07 (0.96 to 1.20) | 1.08 (0.95 to 1.23) | 1.08 (0.94 to 1.23) |
|  | Obesity | 2.16 (2.07 to 2.25) | 1.59 (1.52 to 1.66) | 2.18 (2.03 to 2.35) | 1.44 (1.34 to 1.55) | 2.53 (2.34 to 2.73) | 2.10 (1.95 to 2.27) | 1.27 (1.15 to 1.41) | 1.20 (1.08 to 1.33) | 1.66 (1.50 to 1.83) | 1.46 (1.32 to 1.62) |
|  | Type 2 diabetes | 2.66 (2.51 to 2.81) | 1.31 (1.23 to 1.38) | 2.67 (2.41 to 2.96) | 1.30 (1.17 to 1.44) | 3.21 (2.82 to 3.66) | 1.86 (1.62 to 2.12) | 1.09 (0.98 to 1.22) | 1.07 (0.96 to 1.20) | 1.23 (1.09 to 1.39) | 1.21 (1.07 to 1.36) |
| From diagnosed with COVID-19 to death | Charlson - 1:0 | 12.45 (10.94 to 14.16) | 1.97 (1.72 to 2.25) | 3.93 (2.41 to 6.41) | 1.72 (1.05 to 2.83) | 8.81 (4.38 to 17.72) | 5.39 (2.66 to 10.90) | 1.71 (1.36 to 2.16) | 1.40 (1.11 to 1.77) | 2.56 (2.14 to 3.05) | 2.02 (1.69 to 2.41) |
|  | Charlson - 2:0 | 15.37 (13.47 to 17.54) | 1.60 (1.39 to 1.83) | 10.87 (7.02 to 16.85) | 2.79 (1.77 to 4.40) | 14.27 (7.19 to 28.33) | 5.72 (2.84 to 11.51) | 1.27 (1.01 to 1.61) | 1.12 (0.89 to 1.42) | 1.93 (1.61 to 2.32) | 1.54 (1.28 to 1.85) |
|  | Charlson - 3+:0 | 45.56 (40.68 to 51.02) | 2.34 (2.07 to 2.65) | 24.28 (16.31 to 36.13) | 4.88 (3.18 to 7.49) | 93.56 (51.94 to 168.54) | 28.15 (15.11 to 52.42) | 2.48 (2.04 to 3.00) | 1.85 (1.52 to 2.25) | 3.07 (2.61 to 3.62) | 2.14 (1.82 to 2.52) |
|  | Autoimmune condition | 1.79 (1.59 to 2.03) | 1.10 (0.97 to 1.24) | 3.40 (2.16 to 5.34) | 2.38 (1.52 to 3.74) | 2.10 (1.05 to 4.21) | 1.37 (0.68 to 2.74) | 0.98 (0.79 to 1.22) | 1.03 (0.83 to 1.28) | 0.98 (0.83 to 1.14) | 1.06 (0.90 to 1.24) |
|  | Chronic kidney disease | 12.56 (11.61 to 13.59) | 1.31 (1.21 to 1.42) | 13.35 (8.26 to 21.59) | 2.71 (1.66 to 4.43) | 32.22 (18.03 to 57.59) | 8.23 (4.51 to 15.02) | 1.70 (1.50 to 1.94) | 1.42 (1.25 to 1.62) | 1.55 (1.40 to 1.72) | 1.20 (1.08 to 1.33) |
|  | COPD | 5.12 (4.53 to 5.79) | 1.08 (0.95 to 1.23) | 6.71 (3.94 to 11.41) | 1.47 (0.85 to 2.52) | 15.71 (7.83 to 31.51) | 5.12 (2.54 to 10.32) | 1.09 (0.93 to 1.27) | 1.09 (0.93 to 1.28) | 0.96 (0.75 to 1.22) | 0.96 (0.76 to 1.23) |
|  | Dementia | 20.71 (19.17 to 22.38) | 1.92 (1.77 to 2.08) | NA | NA | NA | NA | 2.63 (2.31 to 2.99) | 2.01 (1.76 to 2.30) | 2.36 (2.14 to 2.61) | 1.86 (1.68 to 2.06) |
|  | Heart disease | 7.05 (6.54 to 7.60) | 1.03 (0.95 to 1.11) | 5.65 (4.01 to 7.95) | 1.78 (1.25 to 2.52) | 5.62 (3.28 to 9.64) | 2.10 (1.21 to 3.65) | 1.19 (1.06 to 1.35) | 1.00 (0.89 to 1.14) | 1.14 (1.03 to 1.26) | 1.02 (0.92 to 1.12) |
|  | Hyperlipidemia | 1.53 (1.38 to 1.70) | 1.01 (0.91 to 1.12) | 1.58 (1.02 to 2.43) | 0.78 (0.51 to 1.21) | 3.62 (2.15 to 6.08) | 1.30 (0.77 to 2.20) | 0.77 (0.64 to 0.92) | 0.94 (0.78 to 1.13) | 0.91 (0.79 to 1.04) | 1.06 (0.92 to 1.22) |
|  | Hypertension | 3.42 (3.16 to 3.70) | 0.95 (0.88 to 1.03) | 2.80 (1.96 to 3.99) | 0.81 (0.56 to 1.16) | 4.35 (2.67 to 7.09) | 1.49 (0.90 to 2.45) | 0.91 (0.80 to 1.03) | 0.92 (0.81 to 1.04) | 1.00 (0.90 to 1.11) | 0.97 (0.87 to 1.07) |
|  | Malignant neoplasm | 5.18 (4.74 to 5.66) | 1.24 (1.13 to 1.35) | 8.08 (5.39 to 12.10) | 1.87 (1.23 to 2.84) | 15.54 (9.78 to 24.69) | 6.34 (3.96 to 10.16) | 1.18 (1.03 to 1.35) | 1.14 (0.99 to 1.30) | 1.20 (1.05 to 1.37) | 1.21 (1.06 to 1.38) |
|  | Obesity | 1.50 (1.38 to 1.64) | 0.98 (0.90 to 1.07) | 2.48 (1.76 to 3.50) | 1.13 (0.80 to 1.59) | 2.80 (1.76 to 4.43) | 1.79 (1.12 to 2.84) | 0.76 (0.65 to 0.87) | 0.91 (0.79 to 1.06) | 0.76 (0.68 to 0.85) | 0.97 (0.87 to 1.09) |
|  | Type 2 diabetes | 5.01 (4.59 to 5.48) | 1.31 (1.20 to 1.43) | 8.70 (6.14 to 12.32) | 2.34 (1.64 to 3.35) | 11.35 (6.75 to 19.08) | 3.67 (2.16 to 6.24) | 1.14 (0.99 to 1.31) | 1.21 (1.05 to 1.40) | 1.20 (1.06 to 1.36) | 1.26 (1.12 to 1.43) |
| From hospitalised with COVID-19 to death | Charlson - 1:0 | 3.26 (2.88 to 3.68) | 1.58 (1.40 to 1.79) | 2.92 (2.18 to 3.89) | 1.98 (1.47 to 2.66) | 2.01 (1.21 to 3.35) | 1.59 (0.95 to 2.64) | 1.49 (1.23 to 1.82) | 1.37 (1.13 to 1.67) | 1.74 (1.41 to 2.15) | 1.45 (1.18 to 1.80) |
|  | Charlson - 2:0 | 3.45 (3.08 to 3.87) | 1.42 (1.26 to 1.61) | 2.69 (1.98 to 3.66) | 1.66 (1.21 to 2.28) | 2.66 (1.70 to 4.15) | 1.83 (1.17 to 2.87) | 1.44 (1.20 to 1.72) | 1.28 (1.07 to 1.54) | 1.51 (1.24 to 1.85) | 1.23 (1.01 to 1.51) |
|  | Charlson - 3+:0 | 6.07 (5.50 to 6.70) | 1.81 (1.62 to 2.01) | 5.20 (3.93 to 6.89) | 3.05 (2.28 to 4.08) | 7.63 (5.14 to 11.31) | 4.77 (3.19 to 7.15) | 1.79 (1.53 to 2.10) | 1.45 (1.24 to 1.71) | 2.30 (1.93 to 2.73) | 1.67 (1.40 to 2.00) |
|  | Autoimmune condition | 1.41 (1.26 to 1.58) | 1.19 (1.06 to 1.33) | 1.88 (1.37 to 2.59) | 1.58 (1.15 to 2.18) | 1.36 (0.82 to 2.25) | 1.13 (0.68 to 1.88) | 1.07 (0.90 to 1.28) | 1.09 (0.91 to 1.30) | 1.20 (1.01 to 1.43) | 1.20 (1.01 to 1.43) |
|  | Chronic kidney disease | 3.10 (2.86 to 3.36) | 1.28 (1.18 to 1.40) | 3.46 (2.53 to 4.74) | 2.17 (1.58 to 2.99) | 4.72 (3.00 to 7.41) | 2.90 (1.83 to 4.58) | 1.41 (1.26 to 1.58) | 1.21 (1.08 to 1.36) | 1.55 (1.36 to 1.77) | 1.22 (1.06 to 1.39) |
|  | COPD | 2.17 (1.95 to 2.42) | 1.15 (1.03 to 1.29) | 3.55 (2.60 to 4.84) | 2.01 (1.47 to 2.76) | 4.05 (2.19 to 7.50) | 2.63 (1.42 to 4.86) | 1.05 (0.91 to 1.20) | 1.03 (0.90 to 1.18) | 1.29 (1.00 to 1.66) | 1.19 (0.92 to 1.53) |
|  | Dementia | 4.14 (3.72 to 4.59) | 1.64 (1.47 to 1.83) | NA | NA | NA | NA | 1.79 (1.51 to 2.11) | 1.47 (1.24 to 1.73) | 2.17 (1.87 to 2.51) | 1.66 (1.43 to 1.94) |
|  | Heart disease | 2.87 (2.67 to 3.09) | 1.24 (1.14 to 1.34) | 2.77 (2.21 to 3.47) | 1.77 (1.40 to 2.23) | 3.29 (2.28 to 4.75) | 2.11 (1.45 to 3.07) | 1.21 (1.09 to 1.35) | 1.10 (0.98 to 1.22) | 1.47 (1.30 to 1.67) | 1.24 (1.09 to 1.40) |
|  | Hyperlipidemia | 1.05 (0.95 to 1.15) | 1.03 (0.93 to 1.13) | 1.15 (0.89 to 1.50) | 0.97 (0.75 to 1.26) | 1.13 (0.75 to 1.71) | 0.76 (0.50 to 1.15) | 1.00 (0.87 to 1.15) | 1.08 (0.95 to 1.24) | 0.94 (0.80 to 1.11) | 1.03 (0.87 to 1.22) |
|  | Hypertension | 1.41 (1.31 to 1.53) | 1.02 (0.94 to 1.10) | 1.54 (1.23 to 1.93) | 1.08 (0.86 to 1.35) | 1.82 (1.30 to 2.56) | 1.19 (0.84 to 1.68) | 0.99 (0.88 to 1.10) | 1.01 (0.90 to 1.12) | 1.00 (0.88 to 1.14) | 1.00 (0.87 to 1.14) |
|  | Malignant neoplasm | 2.21 (2.03 to 2.41) | 1.25 (1.15 to 1.37) | 2.95 (2.25 to 3.88) | 1.83 (1.38 to 2.42) | 2.02 (1.31 to 3.12) | 1.45 (0.94 to 2.25) | 1.27 (1.13 to 1.43) | 1.20 (1.06 to 1.34) | 1.28 (1.09 to 1.49) | 1.21 (1.04 to 1.42) |
|  | Obesity | 1.12 (1.03 to 1.20) | 1.10 (1.02 to 1.18) | 2.06 (1.67 to 2.55) | 1.62 (1.31 to 2.01) | 1.54 (1.12 to 2.11) | 1.36 (0.99 to 1.87) | 0.91 (0.81 to 1.02) | 1.01 (0.90 to 1.14) | 0.87 (0.77 to 0.99) | 1.01 (0.89 to 1.15) |
|  | Type 2 diabetes | 1.76 (1.62 to 1.91) | 1.24 (1.14 to 1.35) | 1.70 (1.31 to 2.20) | 1.17 (0.90 to 1.52) | 3.46 (2.45 to 4.90) | 2.46 (1.73 to 3.50) | 1.10 (0.97 to 1.24) | 1.13 (1.00 to 1.27) | 1.25 (1.08 to 1.44) | 1.30 (1.13 to 1.51) |

**Appendix Figure A4.**
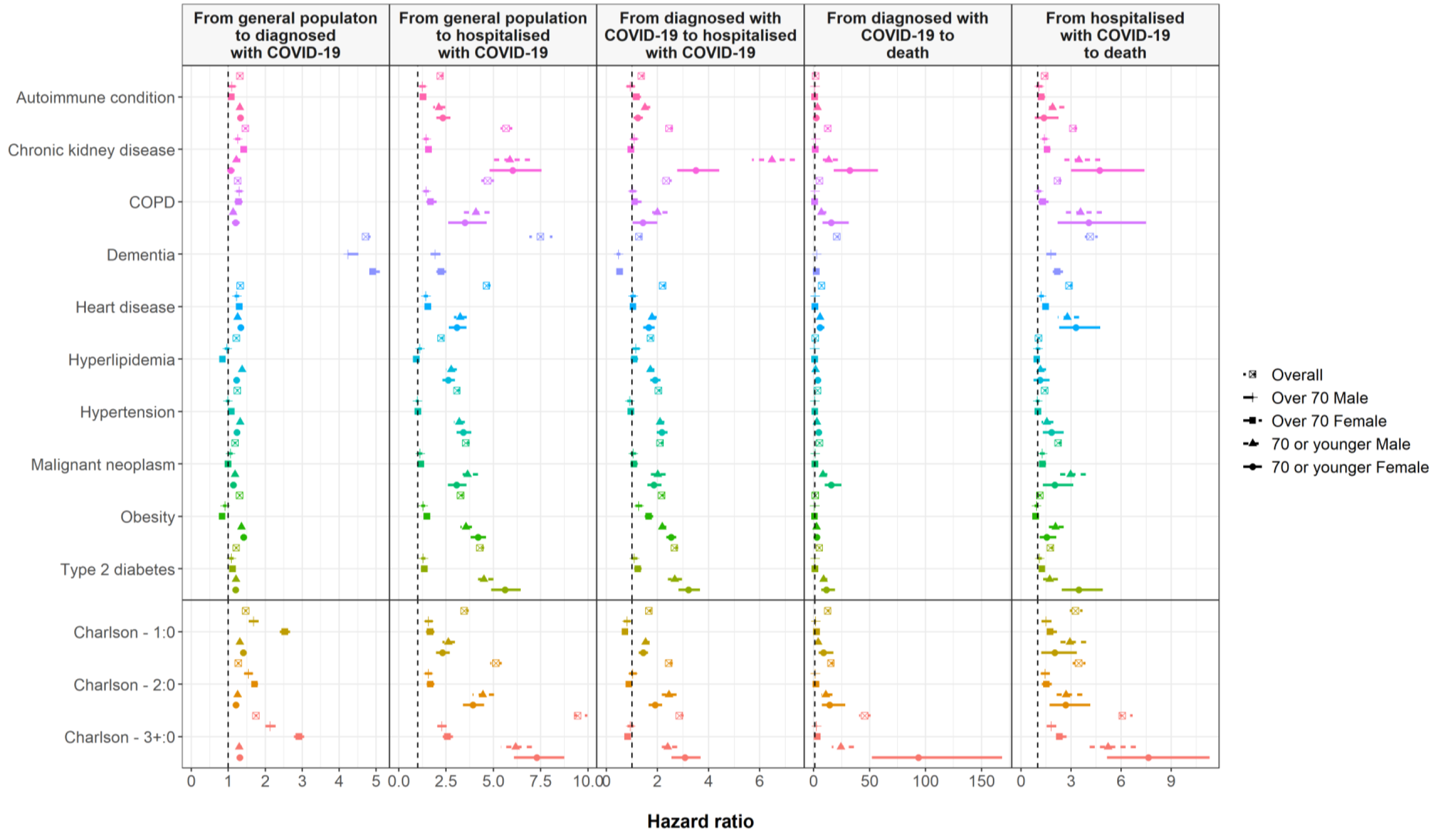
Estimated hazard ratios for comorbidities, unadjusted models

**Appendix Figure A5.**
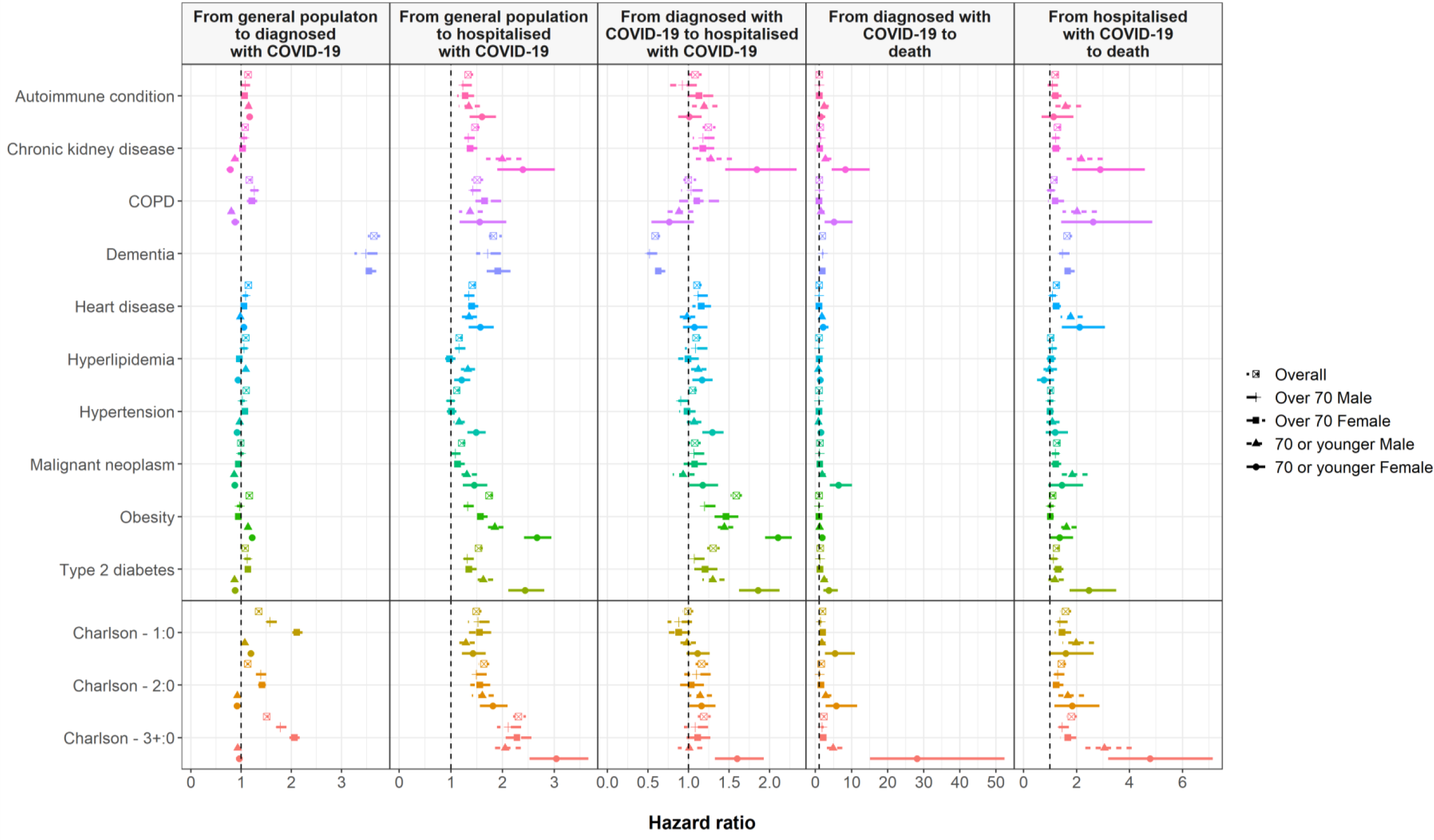
Estimated hazard ratios for comorbidities-models adjusted for age and, when not stratified, by gender

